# Complications and costs to the NHS due to outward medical tourism for elective surgery: a rapid review

**DOI:** 10.1101/2025.04.02.25325086

**Authors:** Clare England, Antonia Needham-Taylor, Nathan Bromham, Juliet Hounsome, Elizabeth Gillen, Jacob Rees Davies, Rhiannon Tudor Edwards, Adrian Edwards, Alison Cooper, Ruth Lewis

## Abstract

Outward medical tourism is when people seek medical treatment in a different country to the one they live in. There are concerns that people travelling abroad for surgery may be at risk of complications when they return home. This review aimed to identify all the studies that describe the impact on the UK NHS of patients who require follow-up care due to outward medical tourism for elective surgery and report on complications, costs and benefits.

The review included evidence available until December 2024. 37 studies were included that described patients who were treated in the NHS for complications arising from elective surgery conducted abroad. 35 were case series or case reports, 19 relating to bariatric surgery, 15 to cosmetic surgery and 1 to ophthalmic surgery. 2 were surveys of plastic surgeons practicing in the NHS. 14 studies included a cost analysis.

The case series and case reports included a total of 655 patients treated by the NHS between 2006 to 2024 for post-operative complications in specific hospitals. Not all studies reported all outcomes of interest. For the studies that reported demographic data, most patients were female (90%). The most common destination for surgery was Turkey (61%). For bariatric surgery tourism, abdominal pain, vomiting, inability to swallow and malnutrition were cited as presenting symptoms, with gastric leak being the most common diagnosis.

Over a third of patients had to have a reversal or revision of the bariatric procedure. For cosmetic surgery tourism, the most common complications were infection and reopening of the surgical wound, with 57% of patients receiving antibiotics. No deaths were reported by any of the studies, although there was evidence that some patients needed complex treatment involving long hospital stays and multiple surgical interventions. Just over a half of patients required at least one investigation or intervention under local or general anaesthetic. Very low certainty of evidence indicates that costs to the NHS from outward medical tourism for elective surgery ranges from £1,058 to £19,549 per patient in 2024 prices. We found no studies that reported on benefits of outward medical tourism.

Awareness-raising campaigns and interventions are warranted to inform members of the public considering going abroad for surgery about the potential for complications. There is a need for a systematic approach to collecting information on the impact on the UK NHS of treating complications arising from outward medical tourism for elective surgery and the associated costs. We still do not know how many people resident in the UK go abroad for elective surgery or how many people subsequently have complications. Without this data we cannot fully understand the amount of risk that people seeking surgery abroad are taking.

**Funding statement:** The authors and their Institutions were funded for this work by the Health and Care Research Wales Evidence Centre, itself funded by Health and Care Research Wales on behalf of Welsh Government.

## 1. BACKGROUND

### 1.1 Who is this review for?

This Rapid Review was conducted as part of the Health and Care Research Wales Evidence Centre Work Programme. The topic was suggested by the Senior Medical Officer for Primary Care, Mental Health, Substance Misuse and Vulnerable Groups Division, and the Deputy Director for Public Health, Improvements and Inequalities, Welsh Government.

This review is intended to inform policy decisions regarding the impact of patients who have received private surgical treatment abroad and require NHS follow-up care, in particular to try avoid unintended consequences and pressures on existing NHS service and limit the impact on health inequity.

### 1.2 Background and purpose of this review

Outward medical tourism is where people seek elective medical treatment in a different country than the one where they live (Hanefeld et al. 2014).It is a practice that has been rising for several decades, and is likely to continue to increase, made attractive by low cost air fares and the use of the internet by medical providers in one country to market their services directly to patients in another (Azlan et al. 2023).

Outward medical tourism potentially creates a problem for health services in the home country, because patients may need post-surgery follow-up at home, and because of the risk of post-surgical complications. Treatment of complications due to outward medical tourism can be costly and made more complicated because full information about the initial surgery may be unavailable. There is limited information on the frequency and type of complications arising from medical tourism. We know that globally, for cosmetic surgery tourism, wound infection and lack of wound healing are the most common complications reported (Alkaelani et al. 2023). We know that complications can be serious and may require treatment in intensive care, further surgery and extensive use of antibiotics. There are reports of multiple organ failure due to sepsis from wound infection and deaths (McCrossan et al. 2021). In 2023, six British nationals were known to have died in Turkey following medical procedures (NaTHNac 2024).

We do not know the scale of the problem that outward medical tourism presents to the UK NHS in 2025. In 2012 Lunt et al. (2014) examined the implications for NHS of inward and outward medical travel, including travel for elective surgery. Medical tourists spanned all ages and were spread across a range of socio-economic groups. The motivations for seeking elective medical treatment outside of the UK were explored qualitatively and found to be variable. A key motivator was availability because the desired procedure was not offered by the NHS, or an individual did not meet NHS eligibility criteria, or NHS waiting times were very long. Another motivator was cost, because private treatment outside of the UK was, or was thought to be, cheaper than private treatment within the UK. It appeared that medical tourists used informal networks to obtain information about treatment abroad and were not well informed about the risks or availability of aftercare by the NHS. Complications arising from medical tourism were found to require NHS treatment, incurring costs, although bariatric surgery tourism was reported to have the potential to result in a saving to the NHS due to the cost-effectiveness of bariatric surgery over time.

In 2010, data from the Office for National Statistics International Passenger Survey indicated that at least 63,000 UK residents travelled abroad for medical treatment (Office for National Statistics 2022). It has been estimated that in 2022 at least 348,000 UK residents travelled abroad for medical treatment (Office for National Statistics 2024) but this is likely to be an underestimate. In 2023, the British Obesity and Metabolic Surgery Society (BOMSS) expressed concern about rising numbers of people experiencing serious complications after weight loss (bariatric) surgery tourism (Aggarwal & Ahmed 2024) and the British Association of Aesthetic Plastic Surgeons advises against travelling abroad for any type of surgery (NaTHNac 2024). More UK-based studies have been published examining complications and associated costs to the NHS. A rapid review, examining all UK-based studies, would inform policy decisions and assist in providing information on potential risks for people considering travelling abroad for medical treatment.

The primary question for this review was:

What are the costs and benefits to the NHS of outward medical tourism for elective surgery? Three secondary questions were considered:

1. What are the long and short-term complications of outward medical tourism for elective surgery treated in the UK by the NHS?
2. What are the costs to the NHS from treatment of complications and follow-up care due to outward medical tourism for elective surgery?
3. What benefits are there to the NHS from outward medical tourism for elective surgery?

Where possible, complications were graded using the Clavien-Dindo classification for postoperative surgical complications (Dindo et al. 2004) (table 1).

**Table 1.**
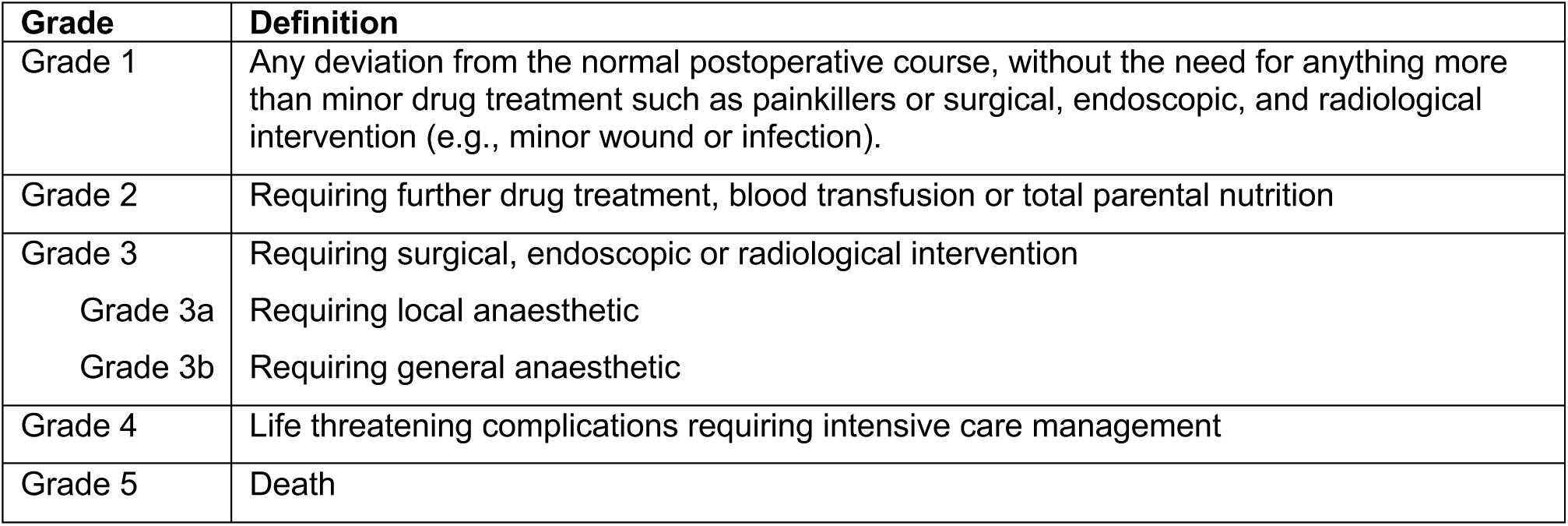
Classification of surgical complications.

## 2. RESULTS

Full details of the study eligibility criteria are described in Section 5, along with the methods used for this review. A detailed summary of the search results and study selection processes, and full description of included studies are summarised in Section 6.

The term ‘initial surgery’ is used to indicate the elective medical tourism surgical procedure that was conducted abroad.

### 2.1 Overview of the Evidence Base

We identified 37 studies for inclusion. All except two studies were conducted in NHS specialist units, or general hospitals and included patients who presented with complications due cosmetic surgery, weight loss (bariatric) surgery or eye (ophthalmic) surgery conducted abroad.

Note: One study, referenced as Burki 2024 is under review and not yet published but some data has been obtained from an article in Lancet Diabetes and Endocrinology.

#### 2.1.1 Bariatric surgery tourism

Nineteen studies, published from 2012 to 2025 focussed on bariatric surgery, of which one also included cosmetic procedures after bariatric surgery. Two studies were published as full peer-reviewed articles (Mandour et al. 2024, Tsang & Jain 2012), sixteen as conference abstracts and findings from another were reported in a commentary (Burki 2024). Twelve studies were case series. For eleven of these, data were obtained retrospectively from medical notes from consecutive eligible patients (Alabdellat et al. 2023, Alnagar et al. 2024, Aly et al. 2022, Beynon et al. 2023, Ftaieh et al. 2023, Gould et al. 2019, Hackney et al. 2022, Hraishawi et al. 2023, Munoz et al. 2021, Patel & Gourgiotis 2024, Burki 2024) and one recruited and followed patients prospectively (Kamel et al. 2017). One of the case series, described in two abstracts, compared emergencies arising from bariatric surgery conducted abroad to emergencies from bariatric surgery conducted in the UK (Alabdellat et al. 2023, Iqbal et al. 2024). The remaining seven studies were case reports, highlighting specific patients of interest (Fakih Gomez et al. 2019, Gan et al. 2017, Mandour et al. 2024, Rashid et al. 2022, Saini et al. 2023a, Saini et al. 2023b, Som et al. 2017, Tsang & Jain 2012). Five of the studies included a cost analysis (Beynon et al. 2023, Gould et al. 2019, Hraishawi et al. 2023, Burki 2024, Munoz et al. 2021). Seventeen of the studies were conducted in sites in England, nine in London hospitals, and two in Northern Ireland. (A detailed description of the studies is provided in Section 6.2. table 10).

#### 2.1.2 Cosmetic surgery tourism

Seventeen studies, published from 2007 to 2024, focussed on cosmetic surgery. Eight studies were published as full peer-reviewed articles (Ahari et al. 2024, Dalmar et al. 2024, Farid et al. 2019, Henry et al. 2021, Jeevan et al. 2011, Martin et al. 2019, Roberts et al. 2024, Thacoor et al. 2019), seven as letters in journals (Bennett et al. 2022, Birch et al. 2007, Long et al. 2021, Sadr et al. 2019, Varma et al. 2022, McCrossan & Jivan 2022) and two as conference abstracts (Segaren et al. 2012, Yoganathan et al. 2023). Twelve studies were case series, either retrospective (Ahari et al. 2024, Bennett et al. 2022, Dalmar et al. 2024, Farid et al. 2019, Henry et al. 2021, Martin et al. 2019, Roberts et al. 2024, Sadr et al. 2019, Segaren et al. 2012, Thacoor et al. 2019, Varma et al. 2022) or unclear (Yoganathan et al. 2023), three studies were case reports (Birch et al. 2007, Long et al. 2021, McCrossan & Jivan 2022) and two were surveys (Birch et al. 2007, Jeevan et al. 2011). Twelve of the case series and case reports were conducted in hospitals in England, two in Northern Ireland, one in Scotland and one in Wales. Eight included a cost analysis (Ahari et al. 2024, Farid et al. 2019, Henry et al. 2021, Martin et al. 2019, Roberts et al. 2024, Sadr et al. 2019, Thacoor et al. 2019, Yoganathan et al. 2023).

Two surveys asked plastic surgeons working in the NHS about patients who had presented with complications from cosmetic surgery conducted abroad. One survey was conducted in the Thames region from April to November 2006 (Birch et al. 2007) and one nationally in 2007 (Jeevan et al. 2011). The surveys did not report demographics, relevant medical history and were imprecise on destination country, complications and treatment. Data from the surveys has not been combined with case series and case reports.

#### 2.1.3 Ophthalmic surgery tourism

One study (Yip et al. 2017) was identified that reported on ophthalmic surgery tourism. This study was published as a full peer-reviewed article and was a case report of five cases seen by one surgeon in England over 10 years and included a cost analysis.

#### 2.1.4 Quality of included studies and assessment of the overall body of evidence

The case reports were generally low to moderate in quality, with one high quality report (Mandour et al. 2024). Lower quality case reports did not report complete demographics, medical history or post-intervention condition. The cost analyses and case series were considered to be at a high risk of bias or included inaccuracies in their results or conclusions. This was primarily because almost all studies were retrospective, with data obtained from medical notes, which can be incomplete or wrongly coded. Most studies included very limited demographic data and very limited previous relevant medical history. It was unclear if data on other outcomes (procedures, complications, treatment or resource use) were complete in most papers, although Farid et al. (2019) reported that six cases were excluded due to lack of follow-up and Yip et al. (2017) reported missing notes for one case. In addition, not all cost analyses were clear about the exact costs included and no study reported whether discounting was applied or not. See section 5.7 for a description of the methods used to for a description of the methods used to assess included studies and section **Error! Reference source not found.**, tables 13, 14 and 15 for the results of the critical appraisal for each study.

The certainty of the evidence on which the findings for the costs (the main focus of the review) has been categorised using the GRADE approach (Grading of Recommendations Assessment, Development and Evaluation) as very low. These ratings provide the degree of confidence we have in the findings, with a high rating indicating, that having assessed the potential problems with the available evidence we are very confident that our summary estimate of the intervention effect represents the true value. Very low certainty indicates that we have very little confidence that our summary of the effect represents the true underlying effect. Further detail on how the overall body of evidence was assessed is provided in section 5.10, and the findings of the assessment in section 6.4 Table 16.

### 2.2 Costs and benefits to the NHS of outward medical tourism for elective surgery

We have considered the three types of elective surgery separately to answer the sub questions:

1. What are the long and short-term complications of outward medical tourism for elective surgery treated in the UK by the NHS?
2. What are the costs to the NHS from treatment of complications and follow-up care due to outward medical tourism for elective surgery?

We were unable to answer sub question 3 since no study examined cost savings or benefits to the NHS from outward medical tourism and no bariatric surgery study reported on long term weight loss or weight maintenance.

#### 2.2.1 Bariatric surgery tourism

##### Patients and procedures

Nineteen studies reported on a total of 385 patients after undergoing bariatric surgery abroad. Twelve studies reported gender and/or age, and no studies reported socioeconomic status or ethnicity. Eight studies reported relevant medical history, ten reported destination country and sixteen reported procedures (see section 6.2 Table 10 for full details of the included studies). The majority of patients were female (83%) and the mean age was 40 years, with a range of 14 to 64 years. Mean pre-operative body mass index (BMI) was 44 kg/m^2^, with a range of 25 to 66 kg/m^2^. BMI at the time of presentation was 37 kg/m^2^ (range 18 to 65 kg/m^2^). The most common destination country was Turkey (62%) and the most common procedures were sleeve gastrectomy (43%) and gastric band (25%) (Table 2).

**Table 2:** Bariatric surgery tourism: combined patient characteristics and procedures.

| **Patient characteristics** |  |
| --- | --- |
| Total number of patients | 385 |
| Gender (n=161) |  |
| Female (n, %) | 134 (83%) |
| Male (n, %) | 10 (6%) |
| Transgender (n, %) | 0 |
| Not reported (n, %) | 17 (11%) |
| Age (n=167) |  |
| Mean (years) | 40.0 |
| Range (years) (n=104) | 14 to 64 |
| Relevant medical history |  |
| Pre-operative BMI (n=42) |  |
| Mean (kg/m<sup>2</sup>) | 44.0 |
| Range (kg/m<sup>2</sup>) | 25 to 66 |
| BMI on presentation (n=58) |  |
| Mean (range) (kg/m<sup>2</sup>) | 37.0 |
| Range (kg/m<sup>2</sup>) | 18 to 65 |
| **Initial surgery** |  |
| Destination countries (n=130) |  |
| Turkey<sup>1</sup> | 80 (62%) |
| Unspecified, excluding Turkey | 45 (35%) |
| Czech Republic | 2 (1.5%) |
| Romania | 2 (1.5%) |
| Thailand | 1 (0.7%) |
| Procedures (n=256) |  |
| Sleeve gastrectomy<sup>2</sup> | 109 (43%) |
| Gastric band | 65 (25%) |
| Gastric bypass<sup>3</sup> | 38 (15%) |
| Gastric balloon | 20 (8%) |
| Post bariatric abdominoplasty | 7 (3%) |
| SASI bipartition | 3 (1%) |
| Revision/conversion | 3 (1%) |
| Gastric plication | 2 (1%) |
| Duodenal switch | 1 (0.4%) |
| Not reported | 8 (3%) |

**Abbreviations and notes:** BMI, body mass index; SASI, Single Anastomosis Sleeve Ileal.
<sup>1</sup>Two case series that did not report specific numbers indicated that Turkey was the most common destination;
<sup>2</sup>Seven cases specified as laparoscopic; <sup>3</sup>Includes Roux-en-y bypass, mini gastric bypass and unspecified gastric bypass.

Two studies collected data on the number of people who required unplanned or emergency care due to complications after bariatric surgery conducted in the UK as well as due to bariatric surgery conducted abroad. Iqbal et al. (2024) reported that 61% of patients received their initial surgery in the UK, 25% of patients abroad, and 14% were unknown. Aly et al. (2022) reported that 80% of patients received initial surgery in the UK and 20% of patients received initial surgery abroad.

##### Question 1: What are the long and short-term complications of outward bariatric surgery tourism treated in the UK by the NHS?

Twelve studies that included 131 patients reported complications in enough detail to combine the data (Alnagar et al. 2024, Fakih Gomez et al. 2019, Gan et al. 2017, Gould et al. 2019, Hraishawi et al. 2023, Kamel et al. 2017, Mandour et al. 2024, Patel & Gourgiotis 2024, Rashid et al. 2022, Saini et al. 2023a, Som et al. 2017, Tsang & Jain 2012). Most studies reporting specific complications included patients that presented with any complication, although two case series specifically recruited patients requiring acute care only (Hraishawi et al. 2023, Patel & Gourgiotis 2024).

Complications were not clearly reported. In many cases symptoms rather than underlying diagnosis were described. The most common presenting symptom was abdominal pain (21%) and the most common diagnosis was gastric leak (17%) (table 3). One study of 41 patients also reported that 4 (10%) patients presented to a tertiary NHS clinic for unspecified routine follow-up care during the study period (Gould et al. 2019).

**Table 3:** Complications due to bariatric surgery tourism.

|  | Number of patients |
| --- | --- |
| **Complications (n=131)** |  |
| **Symptoms** |  |
| Abdominal pain | 28 (21%) |
| Vomiting / reflux | 13 (10%) |
| Malnutrition / nutrition failure | 11 (8%) |
| Unable to swallow | 10 (8%) |
| Weight regain | 8 (6%) |
| Reactive hypoglycaemia | 1 (0.1%) |
| **Diagnosis** |  |
| Gastric leak | 23 (17%) |
| Gastric band erosion | 7 (5%) |
| Stricture / obstruction | 5 (4%) |
| Portal vein thrombosis | 3 (2%) |
| Small bowel ischemia | 2 (1%) |
| Migrated gastric balloon | 2 (1%) |
| Internal hernia | 1 (1%) |
| Unspecified symptoms/diagnosis | 21 (16%) |
| **Clavian-Dindo classification** |  |
| Detailed classification (n=41) |  |
| Grade 1 | 23 (56%) |
| Grade 2 | 5 (12%) |
| Grade 3 | 12 (29%) |
| Grade 4 | 1 (2%) |
| Grade 5 | 0 |
| Overall, grade 3 or above (n=227) | 117 (52%) |
| **Treatment (n=171)** |  |
| Reversal / revision of procedure | 64 (37%) |
| Conversion to another bariatric procedure | 7 (4%) |
| Other surgical treatment | 23 (13%) |
| Non-surgical treatment | 3 (2%) |
| Other (excluding reversal / conversion) | 85 (50%) |

Time since initial surgery was reported by seven studies (n=28) and ranged from 3 to 365 days (Beynon et al. 2023, Gan et al. 2017, Kamel et al. 2017, Mandour et al. 2024, Rashid et al. 2022, Saini et al. 2023b). Studies did not make a clear distinction between short- and longer-term complications.

Detailed Clavian-Dindo classifications were used to report severity of complications by Ftaieh et al. (2023) and it was possible to obtain or derive broad Clavian-Dindo classifications from twelve other studies (Beynon et al. 2023, Fakih Gomez et al. 2019, Gan et al. 2017, Gould et al. 2019, Kamel et al. 2017, Mandour et al. 2024, Munoz et al. 2021, Patel & Gourgiotis 2024, Rashid et al. 2022, Saini et al. 2023a, Som et al. 2017, Tsang & Jain 2012). Overall, at least 117 patients (52%) experienced complications graded 3 or above. There were no deaths reported (Table 3).

Treatment was reported by 13 studies, involving 171 patients (Alnagar et al. 2024, Aly et al. 2022, Fakih Gomez et al. 2019, Gan et al. 2017, Gould et al. 2019, Kamel et al. 2017, Mandour et al. 2024, Munoz et al. 2021, Patel & Gourgiotis 2024, Rashid et al. 2022, Saini et al. 2023a, Som et al. 2017, Tsang & Jain 2012). The most commonly reported treatment was reversal of the procedure, mainly removal of gastric balloons or gastric bands (37%). Only 2% of reported cases received non-surgical treatment, but treatment was not recorded for 50% of cases (Table 3).

##### Question 2: What are the costs to the NHS from treatment of complications and follow-up care due to outward bariatric surgery tourism for elective surgery?

Five studies were included that estimated costs for treatment of complications arising from bariatric surgery tourism (Beynon et al. 2023, Gould et al. 2019, Hraishawi et al. 2023, Burki 2024, Munoz et al. 2021). Studies collected data in 2014 to 2023. It was not clear if all the relevant costs were identified, and the certainty of evidence is very low (see section 2.1.4).

Beynon et al. (2023) reported costs arising from in-patient stays from July 2022 to June 2023. The average cost per patient was reported as £6857.14 (range £600 to £28,625). Gould et al. (2019) reported total treatment costs for patients seen between 2014 to 2019 of £119,920. Clinical appointments accounted for £26,752, investigations for £26,684, hospital admission for £13,920 and costs of revisional surgery for 16 patients were £52,564. Hraishawi et al. (2023) in a study conducted in Northern Ireland, reported that total treatment costs were £737,126, consisting of hospital admission costs of £553,179, cost of non-surgical interventions of £55,607, total cost of scans of £41,728 and surgical intervention costs of £75,413. This study included seven participants with nutritional failure and the costs of dietetic care was estimated at about £17,390. Burki (2024) reported total treatment costs of £560,234 for 35 patients seen in 2022. Munoz et al. (2021) reported the cost of surgical revision or conversion for 32 patients seen between April 2015 to April 2021 as between £4,000 to £7,000 per revision (Table 4).

**Table 4.** Costs due to bariatric surgery tourism.

| Study | Number of patients | Cost year | Description of cost | Original cost as reported | Per patient cost in 2024<sup>2</sup> |
| --- | --- | --- | --- | --- | --- |
| Beynon et al. (2023) | 15 | 2023 | In-patient costs per patient between July 2022 to June 2023 | £6,857.14 (£600-£28,625) | £7,030 |
| Gould et al. (2019)<sup>1</sup> | 41 | 2018 | Total hospital costs between 2014 to 2019 | £119,920 | £3,743 |
| Hraishawi et al. (2023)<sup>1</sup> | 42 | 2022 | Total hospital costs between August 2016 and February 2023 | £737,126 | £19,549 |
| Burki (2024) | 35 | 2022 | Total hospital costs in 2022 | £560,234 | £17,829 |
| Munoz et al. (2021)<sup>1</sup> | 32 | 2020 | Costs per bariatric revision/conversion procedure between April 2015 to April 2021 | £4,000 to £7,000 | £4,986 to £8,726 |

**Note:** <sup>1</sup>Cost year not given, assumed to be the year before publication for calculation of cost in 2024; <sup>2</sup>From the Bank of England Inflation calculator

##### Resource use

Nine studies (200 patients) reported specific use of resources associated with treating complications arising from bariatric surgery tourism (Alabdellat et al. 2023, Beynon et al. 2023, Hackney et al. 2022, Hraishawi et al. 2023, Burki 2024, Mandour et al. 2024, Patel & Gourgiotis 2024, Rashid et al. 2022, Som et al. 2017). All of the nine studies, except Hraishawi et al. (2023) reported length of hospital stay for inpatients. The combined mean length of stay was 17.3 days (159 patients) and the maximum reported stay was 45 days (Patel & Gourgiotis 2024).

Alabdellat et al. (2023) and Iqbal et al. (2024) reported comparisons between patients experiencing complications who had had surgery abroad (n=77) with patients who had had surgery in the UK, either through the NHS or privately (n=188). For patients who had surgery abroad, mean length of stay for the treatment of complications was 20.9 days and median number of stays in intensive care was 8, compared with mean length of stay of 14.4 days and median intensive cares stays of 3 for UK patients (Alabdellat et al. 2023). Post-discharge there appeared to be no difference in subsequent readmissions, diagnostic investigations and interventions between the two groups (Iqbal et al. 2024).

Other resources such as surgery time, number of appointments and number and type of diagnostic tests were not generally reported. Hraishawi et al. (2023) reported that 410 hours of dietetic time were required during the study and Burki (2024) estimated that the overall time needed to treat complications equated to 110 bariatric surgeries (see Section 6.2 Table 10 for other details).

#### 2.2.2 Cosmetic surgery tourism

##### Cosmetic surgery tourism: patients and procedures

The total number of patients reported as having undergone cosmetic surgery abroad by case series or case reports was 265. Twelve studies reported gender and/or age and one reported ethnicity, but no study reported socioeconomic status. Eight studies included some medical history, but reporting was inconsistent. Twelve studies reported destination country and all reported procedures (see Section 6.2 Table 11 for full details). The majority of patients were female (96%) and the mean age was 37 years, with a range of 22 to 59 years. Twenty-eight destination countries were reported, across all continents. The most frequent country was Turkey (62%), with other countries accounting for no more than one to six cases. The most common single procedure was abdominoplasty (25%), although the most common area for surgery was the breast (39%). The average number of procedures per patient was 1.3 (Table 5).

**Table 5:** Cosmetic surgery: combined patient characteristics and procedures.

| **Patient characteristics** |  |
| --- | --- |
| Total number of patients | 265 |
| Gender (n=236) |  |
| Female (n, %) | 227 (96%) |
| Male (n, %) | 5 (2%) |
| Transgender (n, %) | 1 (0.4%) |
| Not reported (n, %) | 3 (1%) |
| Age (n=226) |  |
| Mean (years) | 37 |
| Range (years) (n=77) | 22 to 59 |
| Ethnicity (n=26) |  |
| Asian or Asian British | 11 (42%) |
| White-British | 7 (27%) |
| Black or Black British | 7 (27%) |
| Mixed ethnicity | 1 (4%) |
| **Initial surgery** |  |
| Destination country (n=218) |  |
| Turkey | 136 (62%) |
| Other<sup>1</sup> | 71 (33%) |
| Not recorded | 11 (5%) |
| Procedures (patients=265) (procedures = 355) |  |
| Breast augmentation / enhancement / reduction | 137 (39%) |
| Abdominoplasty (tummy tuck) | 89 (25%) |
| Liposuction | 47 (13%) |
| Gluteal (buttock) augmentation / enhancement | 34 (10%) |
| Facial surgery | 26 (7%) |
| Other body contouring | 14 (4%) |
| Implant exchange | 2 (1%) |
| Gender affirmation (breast removal) | 1 (0.3%) |
| Other | 4 (1%) |
| Unspecified | 1 (0.3%) |
| Average number of procedures per patient | 1.3 |

**Notes:** <sup>1</sup>Lithuania, Czech Republic, Poland, Estonia, Romania, Latvia, Ukraine, Belgium, France, Spain, Brazil, Columbia, Venezuela, Iran, Iraq, Syria, Dubai, Greece, Cyprus, Tunisia, Egypt, USA, Dominican Republic, Pakistan, India, South Korea, Thailand, unspecified – not Turkey.

Three studies also collected data on the number of people who presented with complications due to cosmetic surgery conducted in the UK, either privately or within the NHS, as well as due to cosmetic surgery conducted abroad (Ahari et al. 2024, Bennett et al. 2022, Dalmar et al. 2024). Between 63% and 83% of cases were due to surgery conducted abroad.

In addition, in the surveys, Birch et al. (2007) obtained responses from 35 consultant plastic surgeons working in the NHS, 60% of whom had seen at total of 50 patients with complications due to cosmetic surgery tourism between April to November 2006. Jeevan et al. (2011) received responses from 203 consultant plastic surgeons, working in the NHS, 37% of whom had seen a total of 215 patients with complications or concerns due to cosmetic surgery tourism in 2007. The most common procedures that patients had received abroad in both surveys were breast augmentation/reduction and abdominoplasty.

##### Question 1: What are the long and short-term complications of outward cosmetic surgery tourism treated in the UK by the NHS?

The studies relating to cosmetic surgery tourism reported complications in enough detail to combine the data. The two most commonly experienced complications were infection (45% of patients) and wound dehiscence, where a previously closed surgical incision reopens (43% of patients) (Table 6).

**Table 6.** Complications and treatment associated with cosmetic surgery tourism.

|  | Number of patients (%) |
| --- | --- |
| **Complications (n=265)** |  |
| Infection | 119 (45%) |
| Wound dehiscence | 113 (43%) |
| Haematoma, seroma or abscess | 22 (8%) |
| Necrosis | 20 (8%) |
| Pain | 16 (6%) |
| Implant exposure or rupture | 6 (2%) |
| Removal of drains | 2 (0.1%) |
| Other<sup>1</sup> | 3 (0.1%) |
| Not reported | 32 (12%) |
| **Clavian-Dindo classification** |  |
| **Detailed classification (n=49)** |  |
| Grade 1 | 14 (29%) |
| Grade 2 | 14 (29%) |
| Grade 3 | 16 (33%) |
| Grade 4 | 5 (10%) |
| Grade 5 | 0 (0%) |
| Overall, grade 3 or above (n=139) | 77 (55%) |
| **Treatment of complications (n=169)** |  |
| Antibiotics (oral or intravenous) | 97 (57%) |
| Surgical treatment<sup>2</sup> | 49 (30%) |
| Removal of implants | 14 (8%) |
| Conservative treatment only (dressings, debridement in clinic, suture, drain removal) | 41 (24%) |
| Other<sup>3</sup> | 6 (4%) |
| Not recorded | 22 (13%) |

**Notes:** <sup>1</sup>Nerve injury, abdominal eschar, anaemia requiring transfusion; <sup>2</sup>Surgical treatment numbers includes removal of implants; <sup>3</sup>Ultrasound guided aspiration, blood transfusion, investigation for pulmonary embolism.

Detailed Clavian-Dindo classifications were used to report severity of complications by three studies (Dalmar et al. 2024, Farid et al. 2019, Varma et al. 2022). It was possible to obtain or derive broad Clavian-Dindo classifications from nine other studies (Ahari et al. 2024, Bennett et al. 2022, Birch et al. 2007, Henry et al. 2021, Long et al. 2021, Martin et al. 2019, McCrossan & Jivan 2022, Roberts et al. 2024, Sadr et al. 2019). Overall, at least 77 patients (55%) experienced complications graded 3 or above (Table 6).

Time since initial surgery was reported by seven studies, involving 163 patients (Ahari et al. 2024, Birch et al. 2007, Dalmar et al. 2024, Long et al. 2021, Martin et al. 2019, McCrossan & Jivan 2022, Roberts et al. 2024). All except two patients presented with complications within three months of initial surgery. The remaining two experienced implant rupture over five years post-surgery (Dalmar et al. 2024).

Treatment of complications was not always clearly recorded but most treatment was related to management of infection and wounds. It was possible to combine reported treatment from nine studies, involving 169 patients (Ahari et al. 2024, Bennett et al. 2022, Birch et al. 2007, Henry et al. 2021, Long et al. 2021, Martin et al. 2019, McCrossan & Jivan 2022, Roberts et al. 2024, Thacoor et al. 2019). The most commonly recorded treatment was antibiotics (57% of patients), although it was not always clear as to whether these were oral or intravenous. Thirty percent of patients were recorded as receiving some form of surgical treatment and 24% were treated conservatively with dressings, debridement in clinic or sutures. Fourteen (8%) patients had their implants removed, although three studies reported that one patient each refused implant removal within the NHS and opted to return to the original centre abroad for additional treatment. (Bennett et al. 2022, Martin et al. 2019, McCrossan & Jivan 2022) (Table 6).

The surveys did not report on the specific type of complications. Birch et al. (2007) reported that 53% were emergency admissions and 70% of patients required corrective surgery (Clavian-Dindo classification of 3 or greater). Jeevan et al. (2011) reported that 74% of patients required treatment. Emergency surgery was required for 26% of patients, in-patient non-surgical treatment (e.g. intravenous antibiotics) for 16% of patients and elective surgery for 32% of patients. It was noted that 26% of patients presented with cosmetic dissatisfaction that did not require treatment.

##### Question 2: What are the costs to the NHS from treatment of complications and follow-up care due to outward cosmetic surgery tourism?

Eight studies included a cost analysis (Ahari et al. 2024, Farid et al. 2019, Henry et al. 2021, Martin et al. 2019, Roberts et al. 2024, Sadr et al. 2019, Thacoor et al. 2019, Yoganathan et al. 2023). The studies collected data in 2006 to 2023. All the relevant costs were either not identified, or it was unclear if they were identified, for five studies (Farid et al. 2019, Henry et al. 2021, Martin et al. 2019, Roberts et al. 2024, Yoganathan et al. 2023).

Six studies reported total hospital costs for treatment of complications arising from outward cosmetic surgery tourism for all patients who presented during the study period (Table 7). The studies reported on patients who presented from 2015 to the end of 2023. Roberts et al. (2024), in the largest study conducted in 81 patients, reported total costs of £755,559.68, or £9327.90 per patients, with one patient accounting for £41,2134.33. The largest contributor to the costs was reported to be length of hospital stay at £629,265.00, followed by time spent in theatre at £75,638.09. Ahari et al. (2024) reported total costs of £31,170.80. This was the only study that specified excluded costs, which were those associated with microbiology or pathology, the wages of physicians, and costs to pharmacists and general practices. Farid et al. (2019) reported total costs of £63,803.54. Costs were stratified by Clavien-Dindo classification. Grade 2 accounted for £3,448.60, grade 3 for £18,271.35 and grade 4 for £42,083.59. Henry et al. (2021) reported total costs of £152,946, with an average per patient of £5,882.54 (range £362-£26,585) and the average for a patient requiring surgical intervention of £7,540.37. Martin et al. (2019) reported costs of £23,976.82, with an average per patient cost of £4000 (range £1294-£6291). Sadr et al. (2019) reported costs of £290,456.98. Emergency presentations accounted for £3756.36, elective outpatient appointments for £4007.36, general anaesthesia procedures for £177,773.47, local anaesthesia procedures for £7647.25 and inpatient hospital stay for £97,272. The highest total cost for a single patient, who had complications post-abdominoplasty, was £61,676.

**Table 7.**
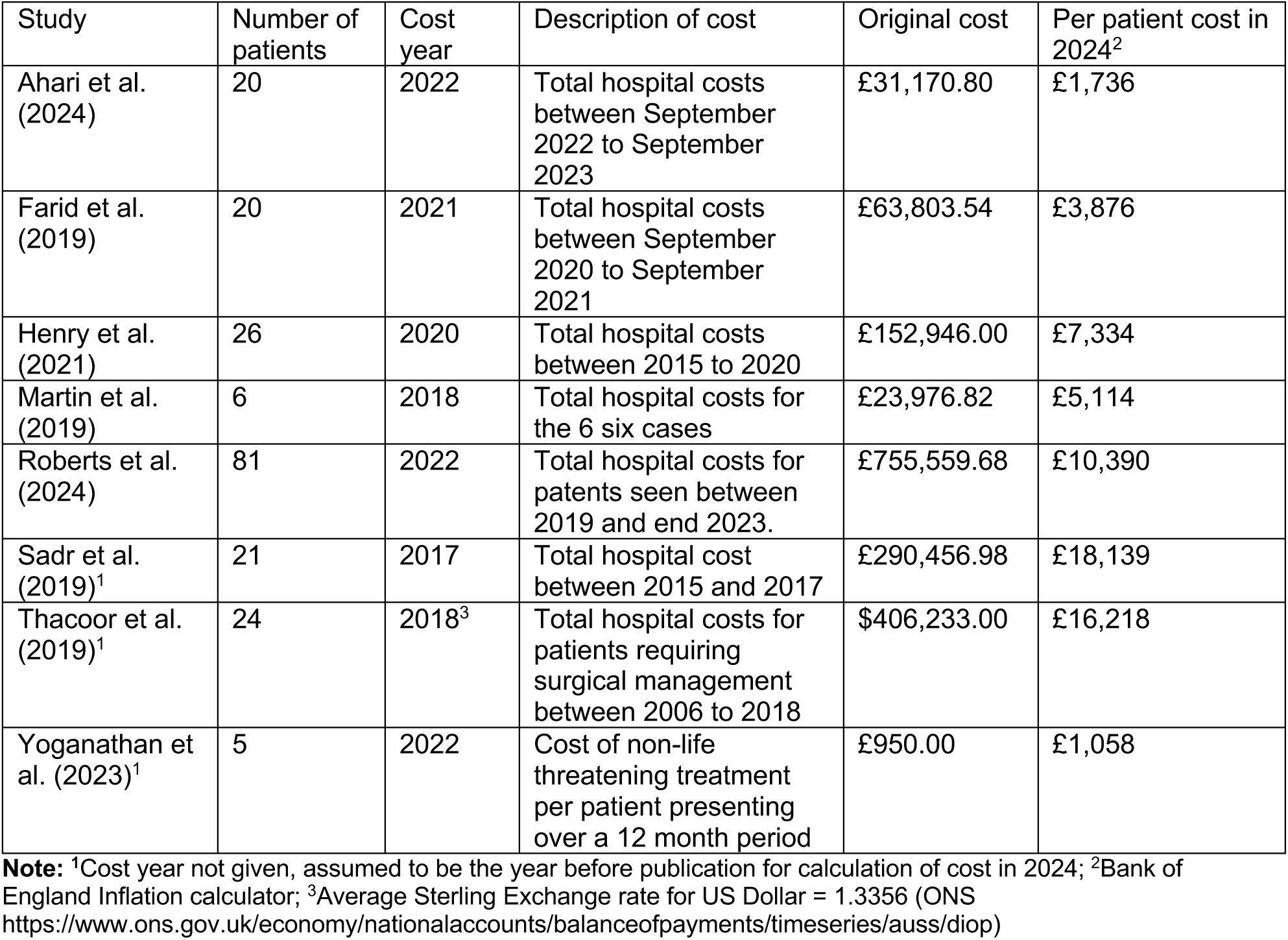
Costs due to cosmetic surgery tourism.

| Study | Number of patients | Cost year | Description of cost | Original cost | Per patient cost in 2024<sup>2</sup> |
| --- | --- | --- | --- | --- | --- |
| Ahari et al. (2024) | 20 | 2022 | Total hospital costs between September 2022 to September 2023 | £31,170.80 | £1,736 |
| Farid et al. (2019) | 20 | 2021 | Total hospital costs between September 2020 to September 2021 | £63,803.54 | £3,876 |
| Henry et al. (2021) | 26 | 2020 | Total hospital costs between 2015 to 2020 | £152,946.00 | £7,334 |
| Martin et al. (2019) | 6 | 2018 | Total hospital costs for the 6 six cases | £23,976.82 | £5,114 |
| Roberts et al. (2024) | 81 | 2022 | Total hospital costs for patents seen between 2019 and end 2023. | £755,559.68 | £10,390 |
| Sadr et al. (2019)<sup>1</sup> | 21 | 2017 | Total hospital cost between 2015 and 2017 | £290,456.98 | £18,139 |
| Thacoor et al. (2019)<sup>1</sup> | 24 | 2018<sup>3</sup> | Total hospital costs for patients requiring surgical management between 2006 to 2018 | \$406,233.00 | £16,218 |
| Yoganathan et al. (2023)<sup>1</sup> | 5 | 2022 | Cost of non-life threatening treatment per patient presenting over a 12 month period | £950.00 | £1,058 |

**Note:** <sup>1</sup>Cost year not given, assumed to be the year before publication for calculation of cost in 2024; <sup>2</sup>Bank of England Inflation calculator; <sup>3</sup>Average Sterling Exchange rate for US Dollar = 1.3356 (ONS https://www.ons.gov.uk/economy/nationalaccounts/balanceofpayments/timeseries/auss/diop)

Thacoor et al. (2019) reported total costs for patients requiring surgical management between 2006 to 2018 of $406,233.00. Yoganathan et al. (2023), in a study conducted in a Welsh breast unit, over an unspecified 12 month period, examined costs relating to non-life threatening care and reported an average of £950.00 per patient (Table 7).

##### Resource use

Eleven studies (225 patients) reported specific use of resources (Ahari et al. 2024, Birch et al. 2007, Farid et al. 2019, Henry et al. 2021, Martin et al. 2019, McCrossan & Jivan 2022, Roberts et al. 2024, Sadr et al. 2019, Segaren et al. 2012, Thacoor et al. 2019, Yoganathan et al. 2023). All studies, except Ahari et al. (2024) and Yoganathan et al. (2023) reported length of hospital stay for inpatients. The combined mean length of stay was 5.9 days (166 patients) and the maximum reported stay was 49 days (Thacoor et al. 2019).

Other resources such as number of day cases, clinic appointments, surgery time and diagnostic tests were reported by no more than one to three studies (see Section 6.2 Table 11 for details). Sadr et al. (2019) was conducted in a tertiary centre specialising in hand trauma and reported that the operative time taken treating complications from 21 cases of cosmetic surgery tourism was comparable to that needed to treat 40 hand-related cases.

#### 2.2.3 Ophthalmic surgery tourism

One paper described brief case reports involving complications after ophthalmic surgery abroad (Yip et al. 2017). All cases were seen by one NHS consultant corneal surgeon over a ten year period (2006 to 2015). A cost analysis was conducted for treatment costs.

##### Ophthalmic surgery: patients and procedures

Five patients, all male, were included. Table 3 shows the reported characteristics and procedures.

**Table 8.**
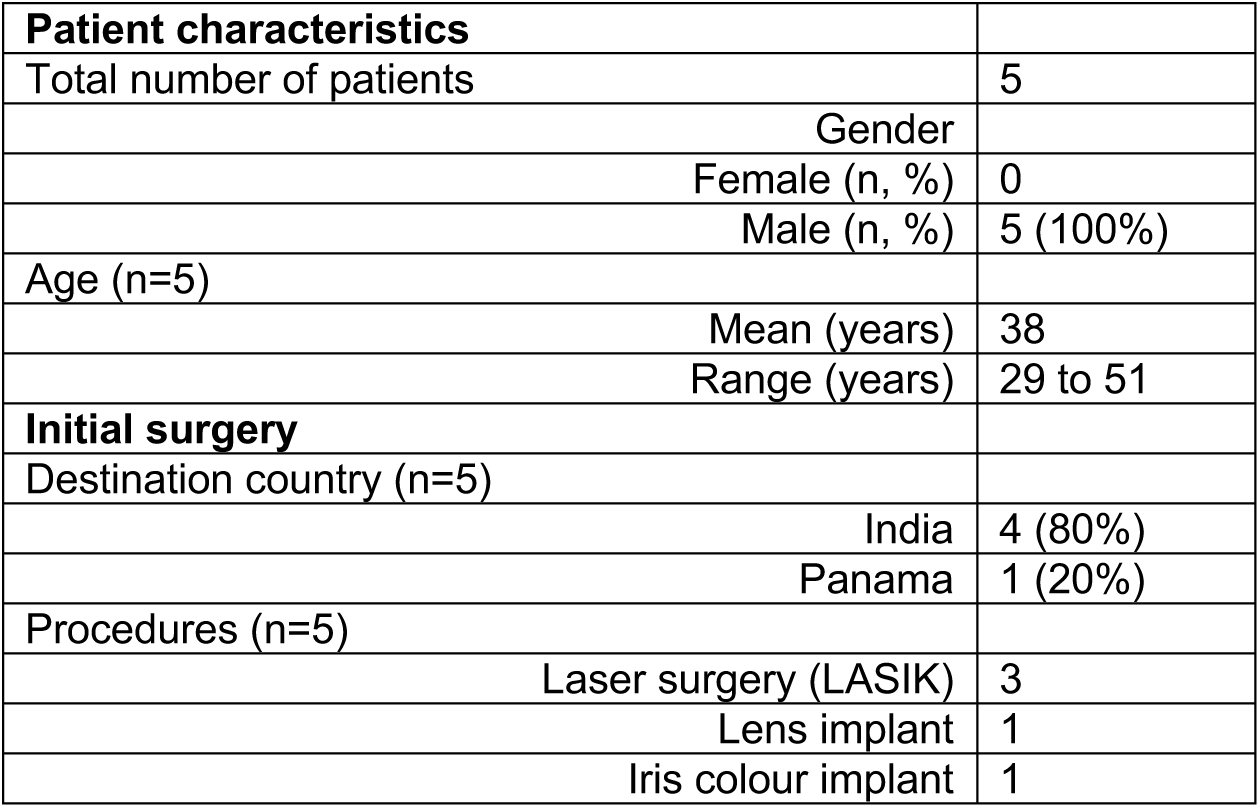
Ophthalmic surgery tourism: combined patient characteristics and procedures.

|  |  |
| --- | --- |
| **Patient characteristics** |  |
| Total number of patients | 5 |
| Gender |  |
| Female (n, %) | 0 |
| Male (n, %) | 5 (100%) |
| Age (n=5) |  |
| Mean (years) | 38 |
| Range (years) | 29 to 51 |
| **Initial surgery** |  |
| Destination country (n=5) |  |
| India | 4 (80%) |
| Panama | 1 (20%) |
| Procedures (n=5) |  |
| Laser surgery (LASIK) | 3 |
| Lens implant | 1 |
| Iris colour implant | 1 |

##### What are the long and short-term complications and costs to the NHS of treatment of outward ophthalmic surgery tourism in the UK?

Four patients presented with reduced sight and one with low grade inflammation between four days to eight months post-surgery. Three patients required topic ointments, lubricants or steroids and two patients required revisional surgery. One patient was diagnosed with a staphylococcus infection and required antibiotics and surgery. Three patients were classified as experiencing grade 2 complications on the Clavian-Dindo score, and two, grade 3a.

Post-treatment two patients experienced long-term mildly reduced sight, one achieved excellent sight and one remained visually impaired and was monitored for further deterioration. The patient with iris colour implants refused advice to remove the implants and continued with daily steroids and ongoing monitoring.

The costs associated with treatment were estimated at £11,583 (cost year, 2016/2017) although some notes were reported to be missing. This is equivalent to £3,039 per patient in 2024 prices. In addition, the costs of eye drops and antibiotics were not included. Over 50 outpatient appointments and four surgical procedures were reported in order to treat the five patients.

### 2.3 Bottom line results for outward medical tourism for elective surgery

Evidence from case series, case reports and surveys show that outward medical tourism for bariatric surgery, cosmetic surgery and ophthalmic surgery can result in serious complications that are treated at specialist units in the UK. Complications arising from bariatric surgery can cause abdominal pain, vomiting, inability to swallow and malnutrition and the most commonly reported treatment is revisional surgery or reversal of the procedure, although non-surgical treatment may not be well recorded. The average length of hospital stay due to bariatric surgery tourism was found to be 17.3 days with a maximum stay of 45 days reported. The most commonly reported complications from cosmetic surgery were infection and reopening of the surgical site which can require antibiotics and surgical treatment under local or general anaesthetic. Conservative management of wounds and infection was also reported, requiring dressings and repeated clinic visits. The average length of hospital stay due to cosmetic surgery tourism was found to be 5.9 days, with a maximum stay of 49 days recorded. Complications from ophthalmic medical tourism can be sight-threatening and treatment can involve multiple appointments.

Costs to the NHS from outward medical tourism for elective surgery ranged from £1,058 to £19,549 per patient in 2024 prices. The certainty of evidence for costs was very low. The highest costs were due to longer stays in hospital and to surgical treatment but it was unclear if all the relevant costs were identified in the majority of studies. Cost and resource use associated with outward medical tourism for elective surgery to GPs and other primary care services were not reported.

## 3. DISCUSSION

### 3.1 Summary of the findings

This rapid review aimed to provide a descriptive summary of the costs and benefits to the NHS of outward medical tourism for elective surgery. The evidence came from case series and case reports conducted in NHS hospitals and specialist units, some of which included an analysis of cost of treatment. We also found two surveys of plastic surgeons working in the NHS, one of which was national, but these were conducted in 2006 and 2007 and did not provide detailed data on most outcomes. We did not identify any studies investigating benefits.

The case series and case reports included a total of 650 patients treated by the NHS between 2011 to 2024 for post-operative complications arising from bariatric or cosmetic surgery abroad and one report of five patients with complications from ophthalmic surgery tourism between 2006 to 2015. We did not identify any studies from the past 10 years that included national or regional level data. Wales was particularly under-represented as we only identified one very low-quality study describing breast surgery tourism that was conducted in a single breast unit (Yoganathan, 2023). This means that case numbers are under-reported, and costs are under-estimated.

Not all studies reported all outcomes of interest. For the studies that reported demographic data, most patients were female (90%), and average age was 38 years (range 14 to 69 years). The most common destination for surgery was Turkey (61%). A wide number of procedures were described, but for bariatric surgery the most common type was sleeve gastrectomy (43%). For cosmetic surgery, the most common single procedure was abdominoplasty (25%), although there was also evidence that patients undergo multiple procedures at the same time.

#### 3.1.1 Question 1: What are the long and short-term complications of outward medical tourism for elective surgery treated in the UK by the NHS?

Some description of specific post-operative complications was available for 401 patients, but reporting was not always clear. Most studies only reported on short-term complications or did not make a distinction between the short and long-term. For bariatric surgery tourism, abdominal pain, vomiting, inability to swallow and malnutrition were cited as presenting symptoms, with gastric leak being the most common diagnosis. Over a third of patients had to have a reversal or revision of the bariatric procedure. For cosmetic surgery tourism, the most common complications were infection and reopening of the surgical wound, with 57% of patients receiving antibiotics.

No deaths were reported by any of the studies, although there was evidence that some patients needed complex and lengthy treatments, with long hospital stays and multiple surgical interventions. Just over half of patients required at least one investigation or intervention under local or general anaesthetic.

#### 3.1.2 Question 2: What are the costs to the NHS from treatment of complications and follow-up care due to outward medical tourism for elective surgery?

Very low certainty of evidence from 14 case series and case reports indicates that costs to the NHS from outward medical tourism for elective surgery ranges from £1,058 to £19,549 per patient in 2024 prices. Data were collected from 2006 to 2023 (the majority over the last 10 years). The highest costs were reported as being related to longer stays in hospital and to surgical treatment, but it was unclear if all the relevant cases and associated costs were identified in the studies.

##### Costs per patient for bariatric, cosmetic and ophthalmic medical tourism

- Total hospital costs/in-patient costs per patient for treatment of complications arising from bariatric surgery complications ranged from £3,743 to £19,549 in 2024 prices (Gould et al. 2019, Hraishawi et al. 2023).
- Costs per revision of bariatric surgery were reported as ranging from £4,968 to £8,729 in 2024 prices (Munoz et al. 2021).
- Total hospital costs per patient for treatment of complications arising from cosmetic surgery tourism ranged from £1,736 to £18,139 in 2024 prices (Ahari et al. 2024, Sadr et al. 2019).
- The cost of non-life-threatening treatment per patient for treatment of complications arising from cosmetic surgery tourism was reported as £1,058 in 2024 prices (Yoganathan et al. 2023).
- The costs of treatment per patient of complications arising from ophthalmic surgery tourism was reported as £3,039 in 2024 prices (Yip et al. 2017).

### 3.2 Limitations of the available evidence

Retrospective case series and case reports are at high risk of bias due to missing information in the records. Most studies did not report on exclusions due to missing data, but there is evidence from two studies that 14% to 23% of potentially eligible cases were excluded for this reason (Alabdellat et al. 2023, Farid et al. 2019). Other studies reported that there was uncertainty in treatment costs due to missing notes (Yip et al. 2017) or an inability to calculate certain patient-specific costs (Roberts et al. 2024). This means that it is likely that the individual studies underestimate both complications arising from medical tourism for elective surgery and associated costs.

More of the evidence for cosmetic surgery tourism came from full peer reviewed articles than for bariatric surgery tourism, which was almost entirely drawn from conference abstracts or letters. Abstracts and letters lack full details on selection of patients and methods of analysis and the reporting of outcomes was often unclear.

Ten included studies were case reports describing between one and five cases in detail. Case reports, as a study design, have limitations for informing practice. The criteria used for selecting cases are seldom stated and cases may be chosen because they are rare or treatment is novel. Case reports can lack generalisability but they do provide a forum to highlight specific issues arising from medical tourism for elective surgery, such as inadequate documentation from the initial surgery, patients travelling shortly after surgery with drains in situ, or patients receiving a type of surgery that would not be considered in the UK.

Two surveys of plastic surgeons were included, one that collected data nationally in 2007 (Birch et al. 2007, Jeevan et al. 2011). However, this survey acknowledged that a limitation was that patients presenting to general medical teams, general breast clinics or other hospitals without a plastic surgery unit would not have been included. In addition, only limited information was collected about complications and treatment.

Five case series reporting on bariatric surgery tourism only included emergency, urgent or acute admissions (Alabdellat et al. 2023, Beynon et al. 2023, Hackney et al. 2022, Burki 2024, Patel & Gourgiotis 2024). This means that these studies did not include patients presenting with non-urgent complications or patients that only required outpatient treatment.

There was some overlap in dates between two studies conducted in the same London hospital (Gould et al. 2019, Munoz et al. 2021). Although the two studies did not include identical outcomes, it is probable that some patients were included in both studies.

NICE recommends that people who have bariatric surgery should be offered follow-up care for a minimum of two years, to include nutritional assessments and dietetic and psychological support (NICE 2025). No study reported on resource use or costs associated with routine long-term follow-up care post bariatric surgery so it is unknown if people who seek bariatric surgery abroad are receiving NHS follow-up care or not.

### 3.3 Strengths and limitations of this Rapid Review

The strength of this rapid review is that it included a systematic and comprehensive search of the available literature, including searching the grey literature and citation searching. The identified studies described place and type of surgery, complications, treatment, use of resources and costs for bariatric and cosmetic surgery tourism. The review includes data from 655 patients who presented with complications due to medical tourism for elective surgery. However, not all outcomes were reported by all studies so each outcome only includes a sub-set of patients.

The main limitation of the review is that it is limited to the cases that have been reported in the literature by researchers who collected data from single specialist units and hospitals, often from patients who presented as emergencies. We only identified one relevant study that presented any national level data and this study only reported on cases observed by NHS plastic surgeons who responded to a survey conducted in 2007. We did not identify any eligible studies conducted in primary care. Only one small low quality study was conducted in Wales. No study has systematically examined complications arising from medical tourism for elective surgery across the UK and the number of UK residents choosing to travel abroad for elective surgery is unknown. This means that the overall complication rate for outward medical tourism for elective surgery in the UK and in Wales is unknown.

The limitations of the available evidence mean it is difficult to tell if there have been changes in the prevalence of medical tourism for elective surgery or changes in the number and type of complications and in treatment costs over time. It is also difficult to make comparisons between bariatric surgery tourism and cosmetic surgery tourism. The reported combined length of hospital stay for bariatric surgery tourism patients was considerably longer than that reported for cosmetic surgery tourism (17 vs 6 days respectively). However, studies reporting on length of stay for bariatric surgery tourism tended to include only emergency or urgent cases, whereas cosmetic surgery tourism studies tended to include both emergency and non-emergency cases. We therefore urge caution in making any direct comparisons. This review does not take into account the social, psychological or economic impact on patients who experience complications from travelling abroad for elective surgery.

### 3.4 Implications for policy and practice

The NHS will continue to provide emergency care to people who experience complications that arise from medical tourism for elective surgery. A UK-wide position is needed on where NHS responsibility lies regarding routine post-operative care and follow up. Should patients be informed that responsibility for ongoing care lies with the original private provider? Would the position vary depending on the type of surgery?

Typing “cosmetic/weight loss surgery in Turkey” into a Google search produces multiple advertisements from clinics offering medical treatment. Roberts et al. (2024) has pointed out that the UK has appropriate regulation to ensure that advertisers are in line with the Advertising Standards Authority (ASA) codes but lacks the ability to enforce them effectively, and the ASA is reliant on the public to report breaches.

Whilst advice and guidance on medical tourism from the NHS and other reputable bodies, does exist, Lunt et al. (2014), commented that people considering elective surgery abroad must seek the advice out themselves. Broad awareness-raising campaigns and interventions are warranted to inform members of the public in Wales considering going abroad for surgery about the potential for complications. It appears that women are more likely to seek elective surgery abroad, especially cosmetic surgery, and an information campaign should be targeted appropriately. Lunt et al. (2014) also advised that consideration should be given to the best way to disseminate information to potential medical tourists. It may be that health professionals with responsibility for informing people as to the availability of treatment in the UK would be well-placed to discuss the implications of seeking treatment abroad.

Content of an awareness campaign could include that for people who require treatment for complications, this often entails treatment needing anaesthetic, and for a significant number of people, may require reversal of the procedure they had abroad. For example, reversal of weight loss surgery or removal of cosmetic implants.

People seeking medical treatment abroad should be made aware of what the NHS will be responsible for in terms of treating complications and the scale of the costs for non-emergency treatment for which they may become personally liable.

Consideration by Government is warranted about the value of requiring insurance to cover costs of potential complications for people going abroad for surgical procedures. Consideration should be given as to whether it is worth attempting to recoup costs from surgical providers abroad.

A driver for bariatric surgery tourism is that weight management services in the UK are inadequate to meet need. In England, 37 of 42 integrated care boards have no policy for treatment of obesity and seven have closed specialist weight management services despite the number of referrals to these services increasing, leading to longer treatment waiting times (Byrne 2024).There is a need to ensure access to weight management services, including weight loss surgery for people who meet NHS criteria, is improved.

### 3.5 Implications for future research

We still do not know how many people go abroad for elective surgery or how many people subsequently have complications. Lunt et al. (2014) advised that robust, reliable data about who is travelling abroad for elective surgery, where they are travelling to and for what medical procedure is needed. Without this data we cannot fully understand the amount of risk that people seeking surgery abroad are taking.

There remains a need to examine the full extent of the impact on the UK NHS of treating complications arising from outward medical tourism for elective surgery and the associated costs. At present the available evidence comes from small studies conducted almost entirely in specialist plastic or bariatric services by local researchers. The scale of the problem in Wales is almost completely unknown. There is a need for a systematic approach to collecting data across the UK, or the devolved nations, that includes other specialities such as orthopaedics. In addition, we did not identify any studies that were conducted in primary care so we do not know what the impact of outward medical tourism for elective surgery is on GP and community services. We do not know if people who have had surgery abroad are seeking non-emergency follow-up care from their GPs. A prospective registry of cases may assist in quantifying the problem for both primary and secondary care.

Future studies should collect and report full demographic details including gender, ethnicity and socioeconomic status. The reporting of Clavian-Dindo classification is a useful way to compare severity of complications across different types of surgery.

There is no evidence comparing the short or long-term health of people who travel abroad for elective surgery with those treated in the UK (either in the NHS or privately). Lunt et al. (2014) advised that direct comparison would allow for a better estimate of costs and benefits to the NHS and provide information on longer term social and economic impact on patients.

### 3.6 Economic considerations^*^

- The economic impact of outward medical tourism for the NHS was thought to be highly variable as medical tourists formed a heterogenous group, across ages, genders, and socioeconomic backgrounds.. (Parliamentary Office of Science and Technology 2020). However, the evidence identified in this review suggested women are more likely to seek elective surgery abroad, especially cosmetic surgery. Further, no studies assessed the impact socioeconomic status had on the likelihood of seeking elective surgery abroad.
- While being one motivating factor, cost alone is never a sole motivator and often not the primary motivation for UK patients seeking treatment abroad (Lunt et al. 2014).
- Rectification surgery and follow-up care following elective care abroad place financial and material pressures on the NHS. The extent to which insurance could mitigate these is currently unclear.

* *This section has been completed by the Centre for Health Economics & Medicines Evaluation (CHEME), Bangor University*

## Data Availability

All data produced in the present study are available upon reasonable request to the authors

## Abbreviations

Acronym: Full Description
BMI: Body mass index
GRADE: Grading of Recommendations Assessment, Development and Evaluation
NHS: National Health Service
NICE: National Institute of Health and Care Excellence
ONS: Office for National Statistics

## Glossary

Abdominoplasty: (tummy tuck) is cosmetic surgery to change the shape of the abdomen, typically removing excess loose skin, fat and stretch marks around the abdomen.
Bariatric surgery: is surgery that makes the stomach smaller to promote weight loss.
Breast augmentation or enlargement: is surgery to increase the size of breasts or change their shape.
Gastric leak: is when digestive juices and partially digestive food leaks thorough a surgical join in the stomach.
Hematoma: is a collection of blood in a wound.
Liposuction: is a cosmetic procedure to remove fat from specific areas of the body.
Necrosis: is the death of tissues of the body.
Seroma: is a collection or pocket of clear fluid in the wound.
Wound dehiscence: is when a surgical wound reopens after it has been closed.

## 5. RAPID REVIEW METHODS

### 5.1 Eligibility criteria

**Table 9:** Eligibility criteria.

|  | Inclusion criteria | Exclusion criteria |
| --- | --- | --- |
| **Setting** | Any NHS setting in the UK. |  |
| **Population** | People who are normally resident in the UK<br>Aged 18+. |  |
| **Exposure** | Elective surgery conducted outside of the UK that is not funded by the NHS (known as elective surgery tourism). | • Emergency surgery<br>• Semi-urgent surgery<br>• Transplant surgery<br>• Cancer treatment<br>• Fertility treatment<br>• Dental treatment<br>• Private surgery within the UK<br>• Treatment outside the UK funded by the NHS |
| **Comparison** | None or elective surgery conducted in the UK by the NHS |  |
| **Outcome measures** | *Primary outcomes(s):*<br>Observed or calculated costs to the NHS in treating patients presenting with complications from elective surgery tourism.<br>Observed or calculated savings to the NHS from elective surgery tourism.<br>*Secondary outcomes(s):*<br>Type and frequency of elective surgery tourism<br>Demographics (includes age, gender, ethnicity, socioeconomic status) of people undertaking elective surgery tourism.<br>Procedure(s) reported.<br>Short and long-term complications.<br>Treatment of complications.<br>Other follow-up care provided by the NHS.<br>Observed or calculated waiting times for elective surgery.<br>Use of NHS resources. |  |
| **Study design** | All quantitative study types including case reports, case series and retrospective and prospective studies.<br>Studies reporting any type of economic evaluation, partial economic evaluation, cost-of-illness analysis or costing studies will be included. | Opinion pieces, editorials, and commentaries.<br>Qualitative studies<br>Minutes of meetings |
| Language of publication | English |  |
| Publication date | After March 2012 |  |
| Publication type | Peer reviewed studies, pre-prints, conference abstracts, letters, grey literature (inc. dissertations, academic, third sector and industry reports, government documents, policy statements). |  |
| Publication date | After March 2012 |  |
| Other factors | Studies that include data from people who received surgery privately in the UK and from people who received surgery abroad will only be included if the data can be disaggregated for costs and / or complications. |  |

#### Definitions

- Elective surgery refers to operations that are planned in advance.
- Emergency surgery refers to operations that must be performed immediately, typically within 24 hours of admission.
- Semi-urgent surgery is a term that is sometimes used for life-preserving surgery that does not need to be performed immediately but should not be delayed too long. It is also known as semi-elective or limited term surgery.
- Costs include medicine costs, administration and monitoring, other health service resource use costs associated with the managing the disease (e.g. GP visits, hospital admissions), and costs of managing adverse events caused by treatment.
- Benefits to the NHS will be measured through calculated or observed savings, a reduction in resource use or reduction in overall waiting times for elective surgery.

### 5.2 Reasons for exclusion

The purpose of this rapid review is to inform policy in areas where there are uncertainties about responsibility for the costs of treatment of complications or managing follow-up. Policy is clear that follow-up care will be offered following fertility treatment, whether received in the UK or abroad. Non-emergency follow-up care or revisions following dental tourism are not covered as a part of NHS dental treatment (NHS England 2024). Fertility treatment and dental treatment abroad will therefore be excluded. In common with Lunt et al. (2014) transplant surgery tourism will also be excluded. There is a separate body of literature that examines transplant surgery tourism, which has specific ethical dimensions.

### 5.3 Incorporating data from existing reviews

Lunt et al. (2014) searched up until March 2012 and was used to identify eligible primary studies published before 2012. Any relevant systematic, rapid, or scoping reviews published after 2012 was checked for eligible primary studies.

### 5.4 Literature search

MEDLINE (Ovid), Embase, Cinahl, Scopus, the Cochrane Library, Proquest (theses) were searched on the 19^th^ November 2024, limited to between 2012 and the 19^th^ November 2024. Overton and Open Alex were searched on the 06/12/2024. Other grey literature sources were searched on the 11^th^ December 2024. See Section 8 (Appendix: search strategies for full details).

### 5.5 Study selection process

Using Endnote ^TM^ (Thomson Reuters, CA, USA), two reviewers dual-screened 10% of titles and abstracts independently. Agreement exceeded the 80% agreement threshold set out in the protocol, so the remaining titles and abstracts were screened by the primary reviewer alone. Following this, 20% of all full texts were dual screened and agreement exceeded the 80% threshold so the remaining records were screened by the primary reviewer alone. During independent screening, the primary reviewer consulted with the secondary reviewer in the case of any uncertainties. We had initially planned to only include studies conducted in over 18s, as per table 9, but not all relevant studies reported age or included an age restriction, so we removed this criteria.

### 5.6 Data extraction

The following data was extracted where available:

- Study information, including author, year, study design, study setting, data collection period, inclusion and exclusion criteria.
- Details of participants, including number and demographics (age, gender, ethnicity, socioeconomic status, medical history relevant to elective surgery e.g. for bariatric surgery, weight before surgery)
- Destination country
- Number and type of elective surgical procedures reported
- Complication-related outcomes, including number, type, and treatment
- Any other follow-up care provided
- Use of NHS resources
- Impact on waiting lists
- Costs from treating complications or providing follow-up care
- Cost-savings to the NHS from elective surgery tourism

Data was extracted by a single reviewer and quality assured by a second reviewer.

### 5.7 Quality appraisal

Study quality was assessed using the appropriate JBI critical appraisal tools selected for each included study design (e.g., Checklist for Case Series, Checklist for Economic Evaluations). Critical appraisal was completed by individual reviewers and checked by a second reviewer. Studies of all quality were included.

### 5.8 Synthesis

Due to the heterogeneity of the outcomes and costs a narrative synthesis was undertaken. The evidence was grouped by type of elective surgery and outcome and used the sub questions as a framework.

### 5.9 Assessment of body of evidence

Certainty in the overall body of evidence was assessed using GRADE for the primary outcome measure of cost. No study reported on any benefits of outward medical tourism for elective surgery.

## 6. EVIDENCE

### 6.1 Search results and study selection

**Figure 1:**
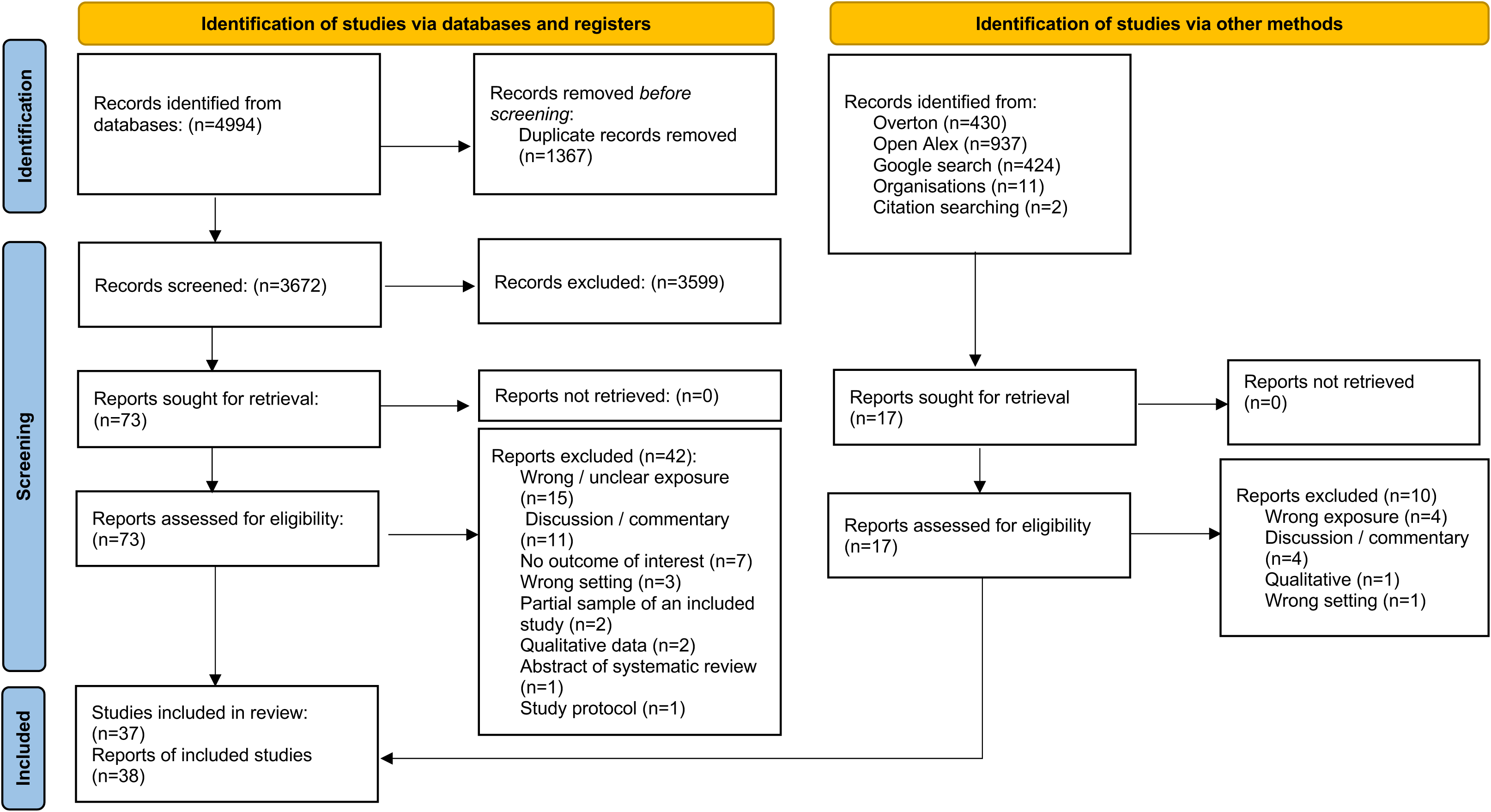
PRISMA flow diagram.

### 6.2 Data extraction - detailed summary of included studies

Studies related to bariatric surgery tourism are summarised in table 10, those related to cosmetic surgery tourism are summarised in table 11 and the study relating to ophthalmic surgery tourism are summarised in table 12.

**Table 10:**
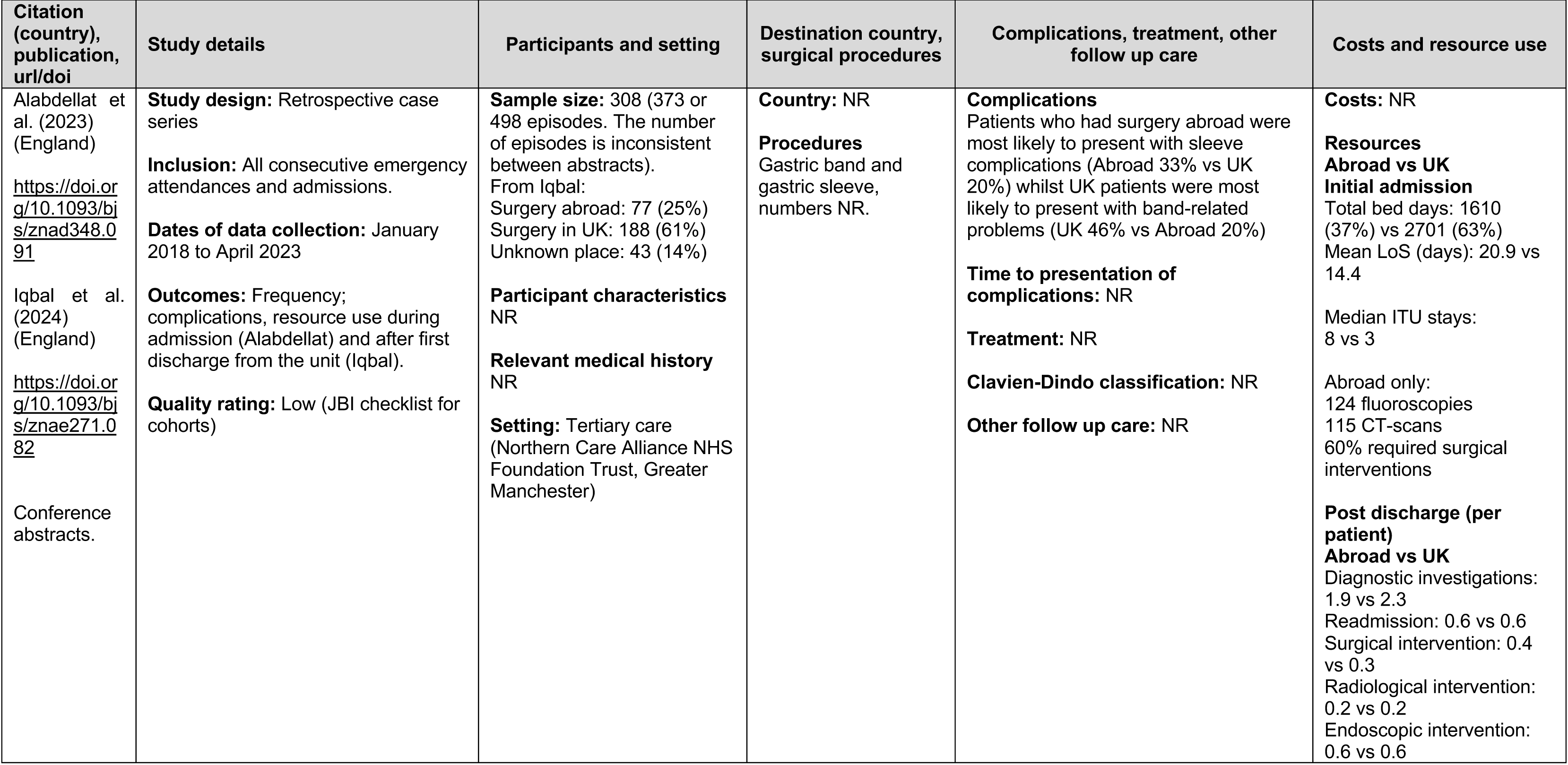
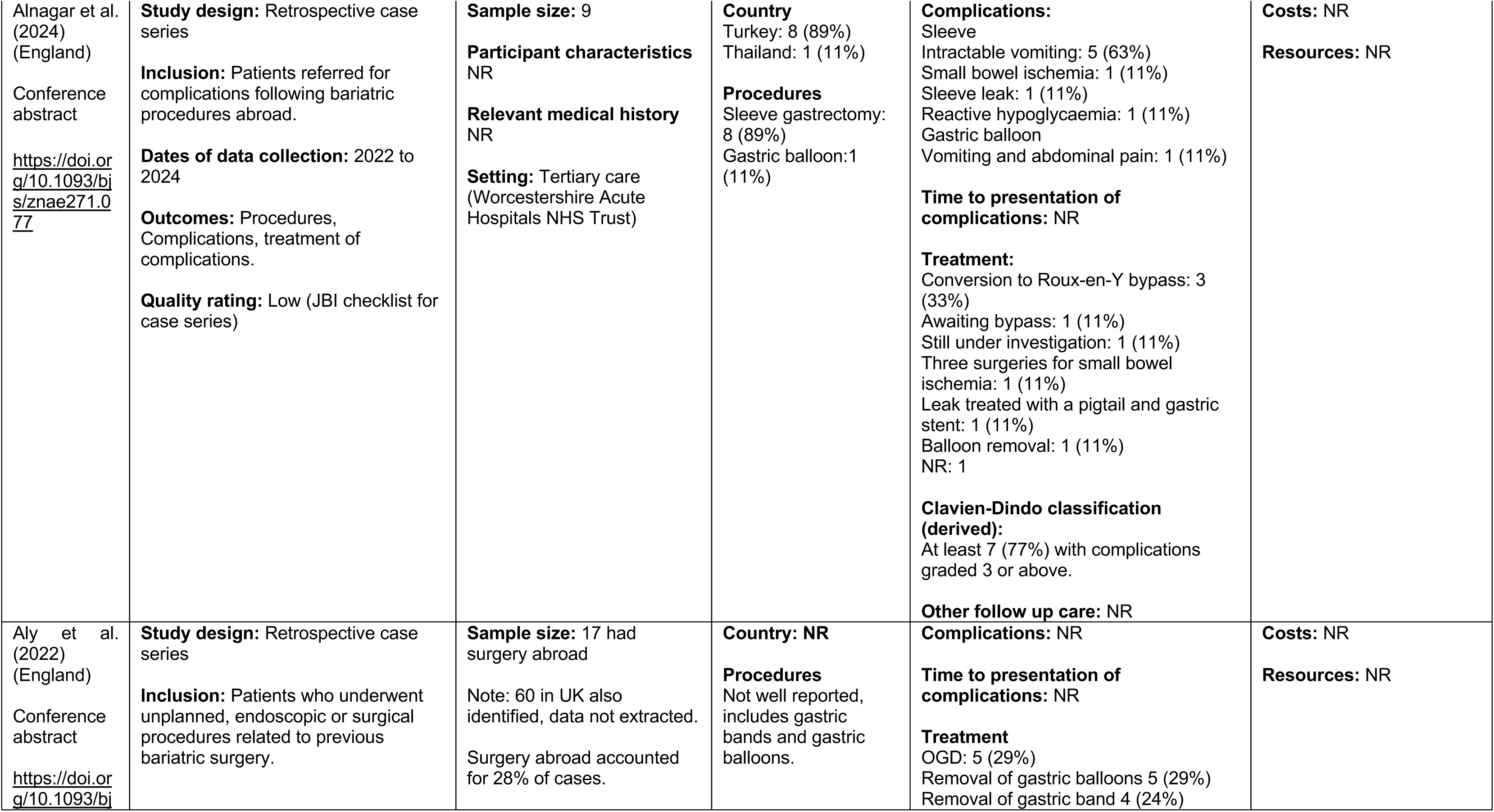
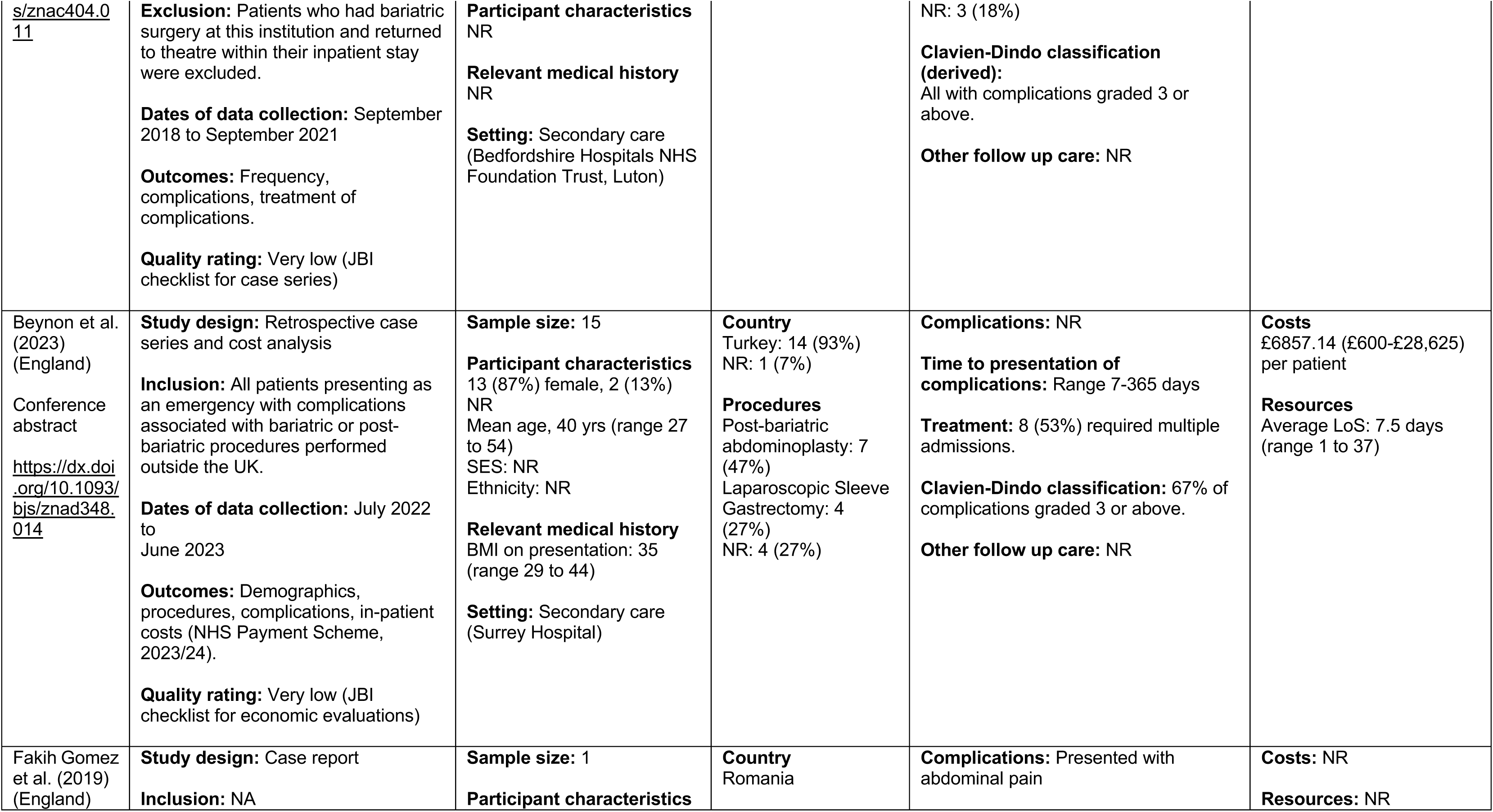
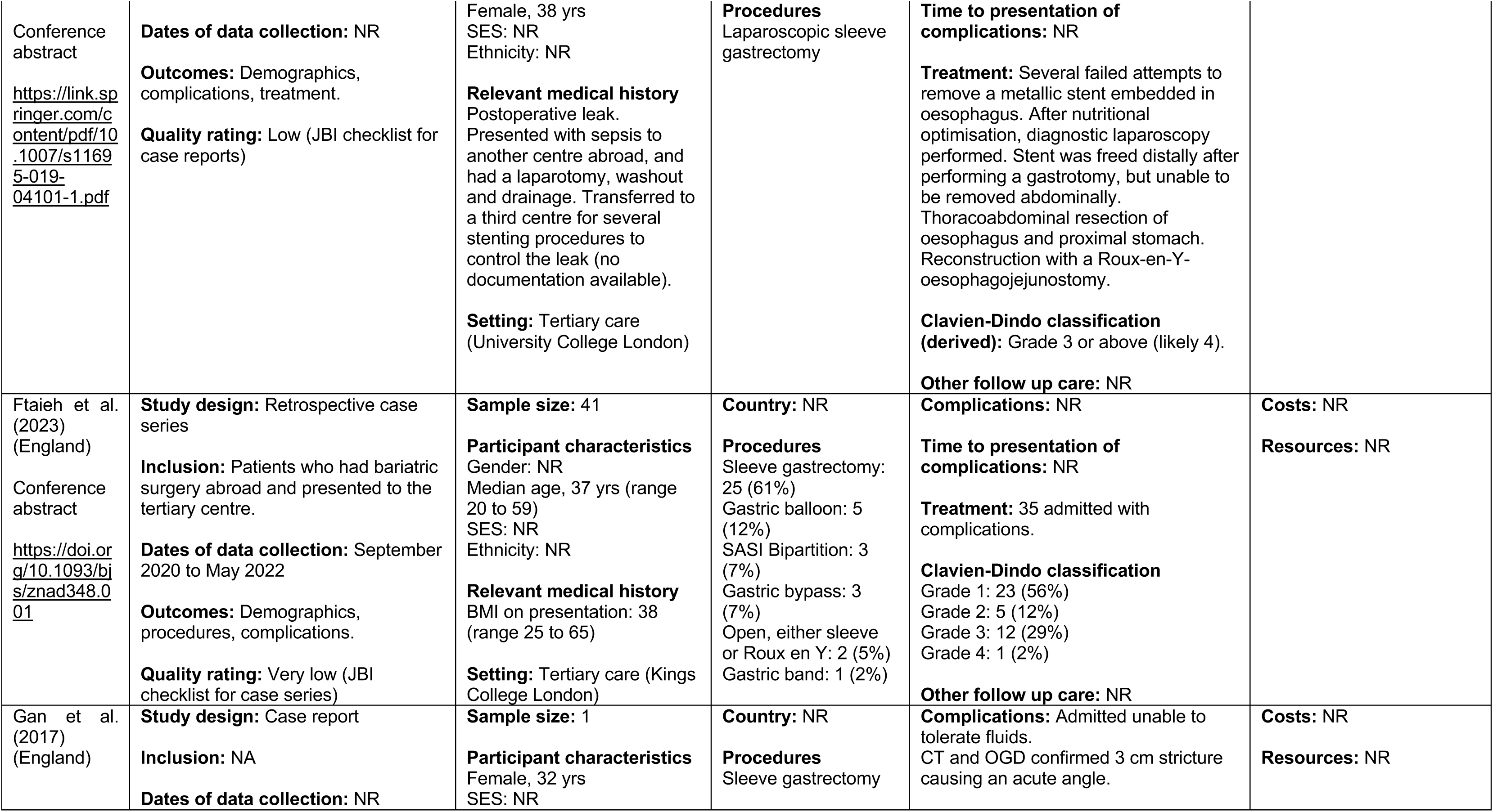
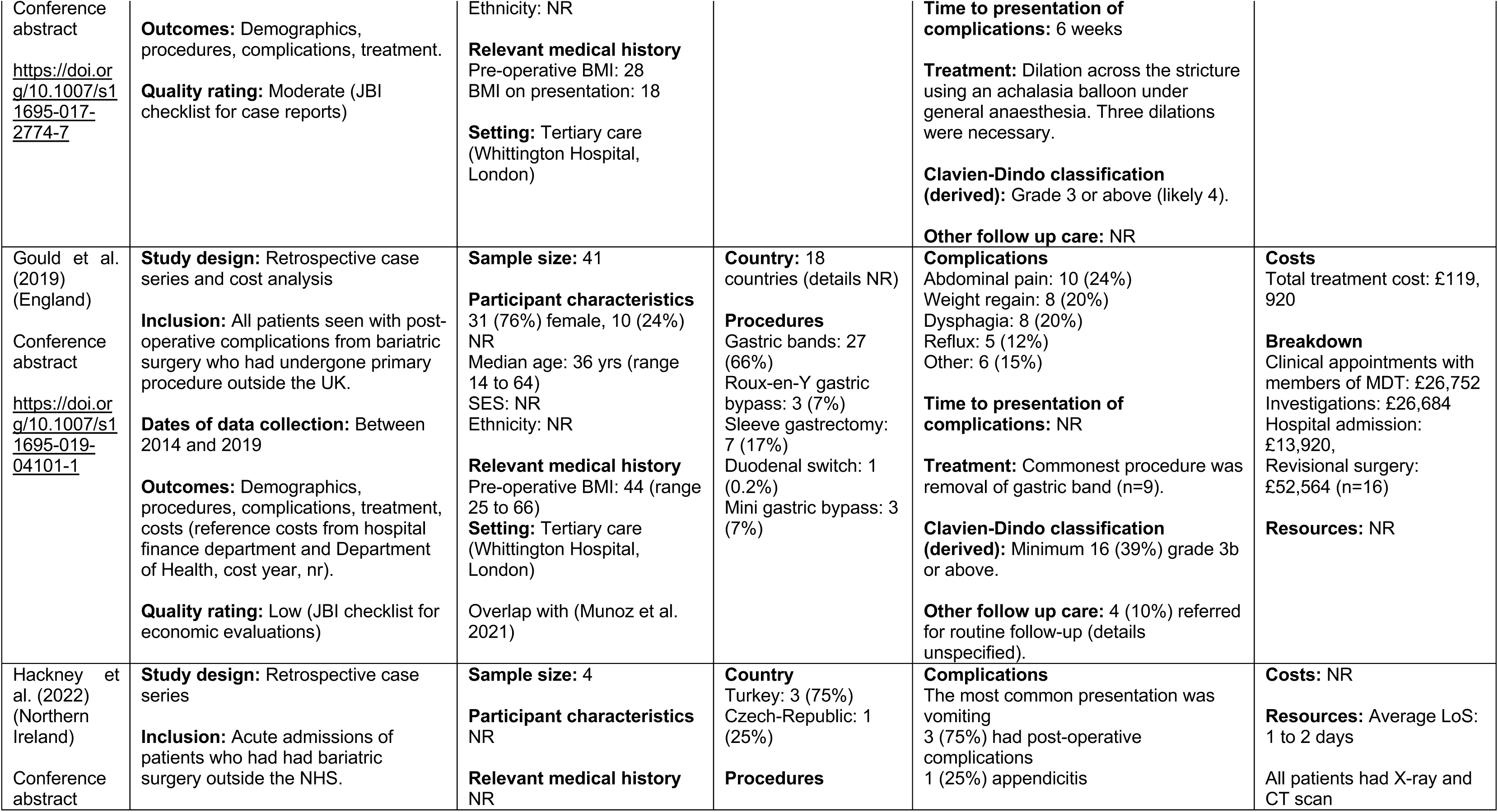
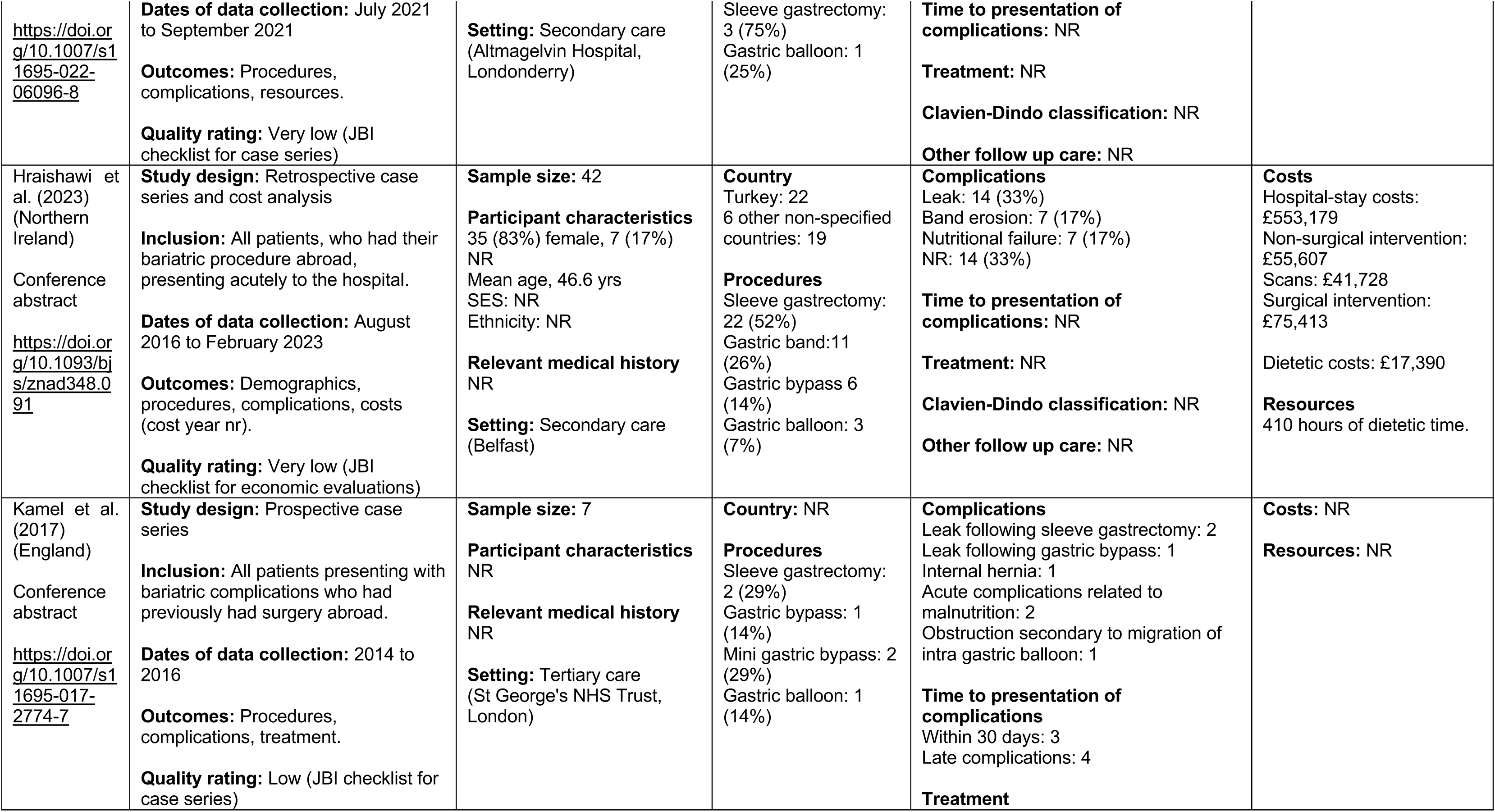
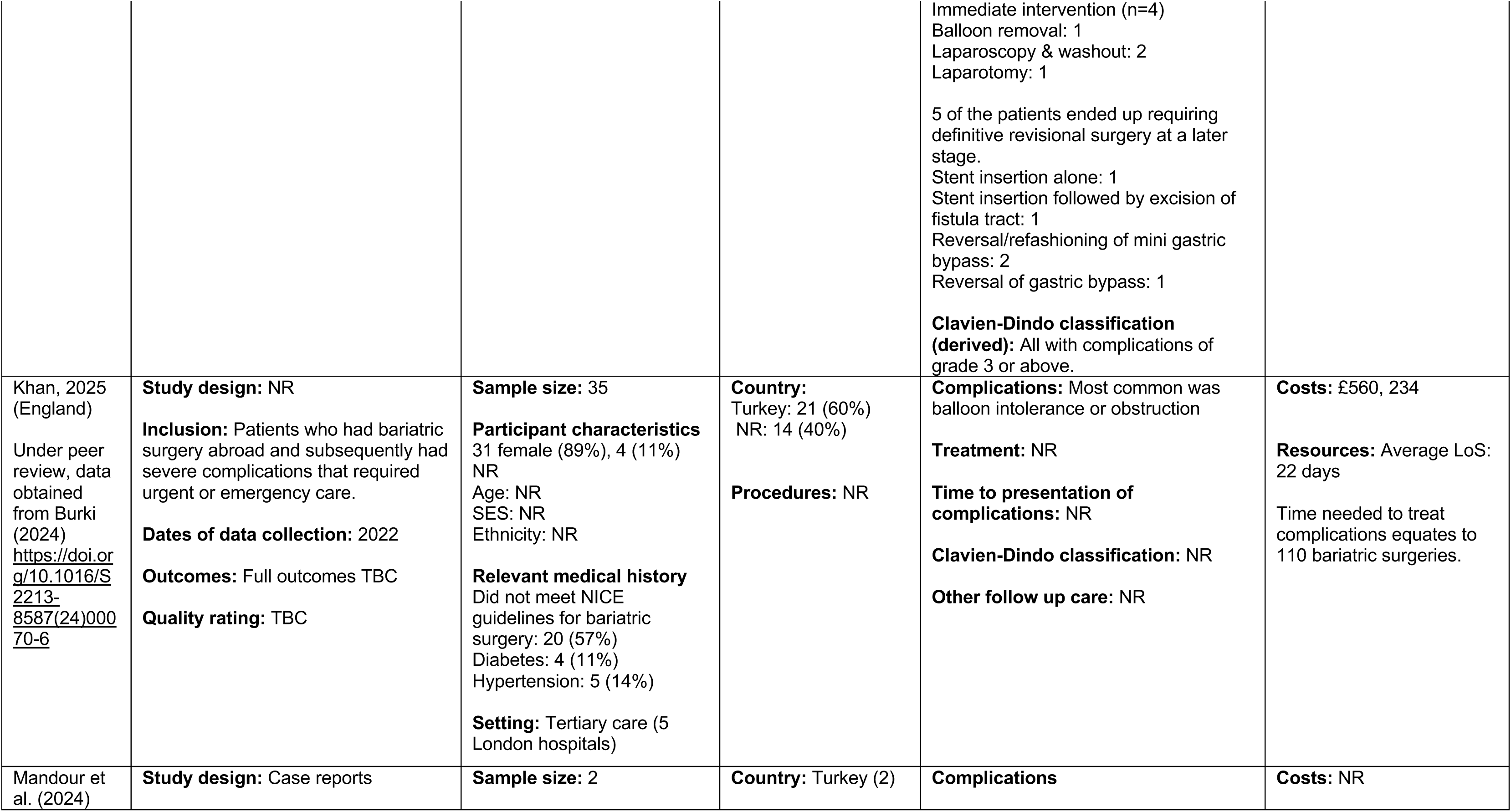
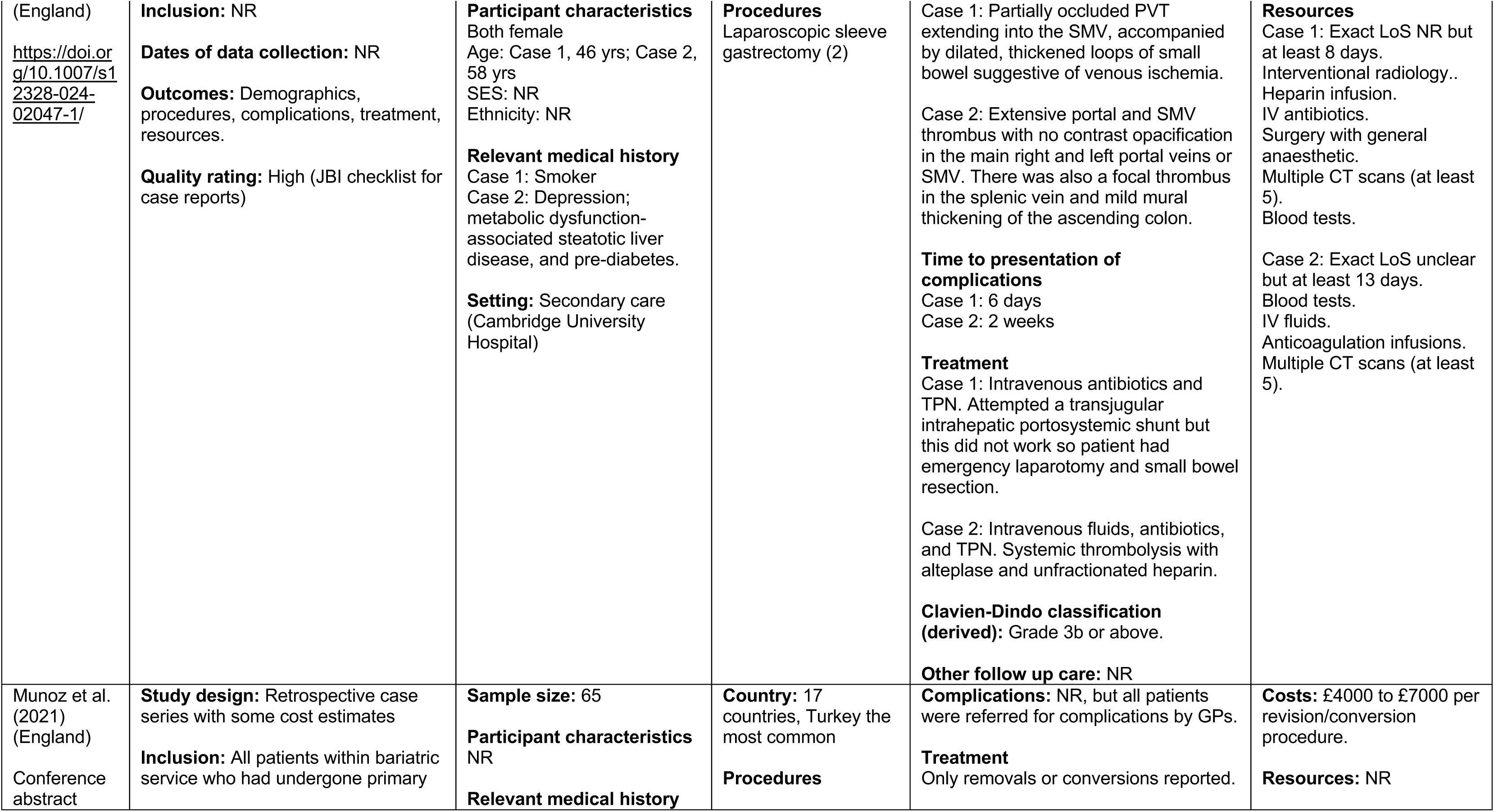
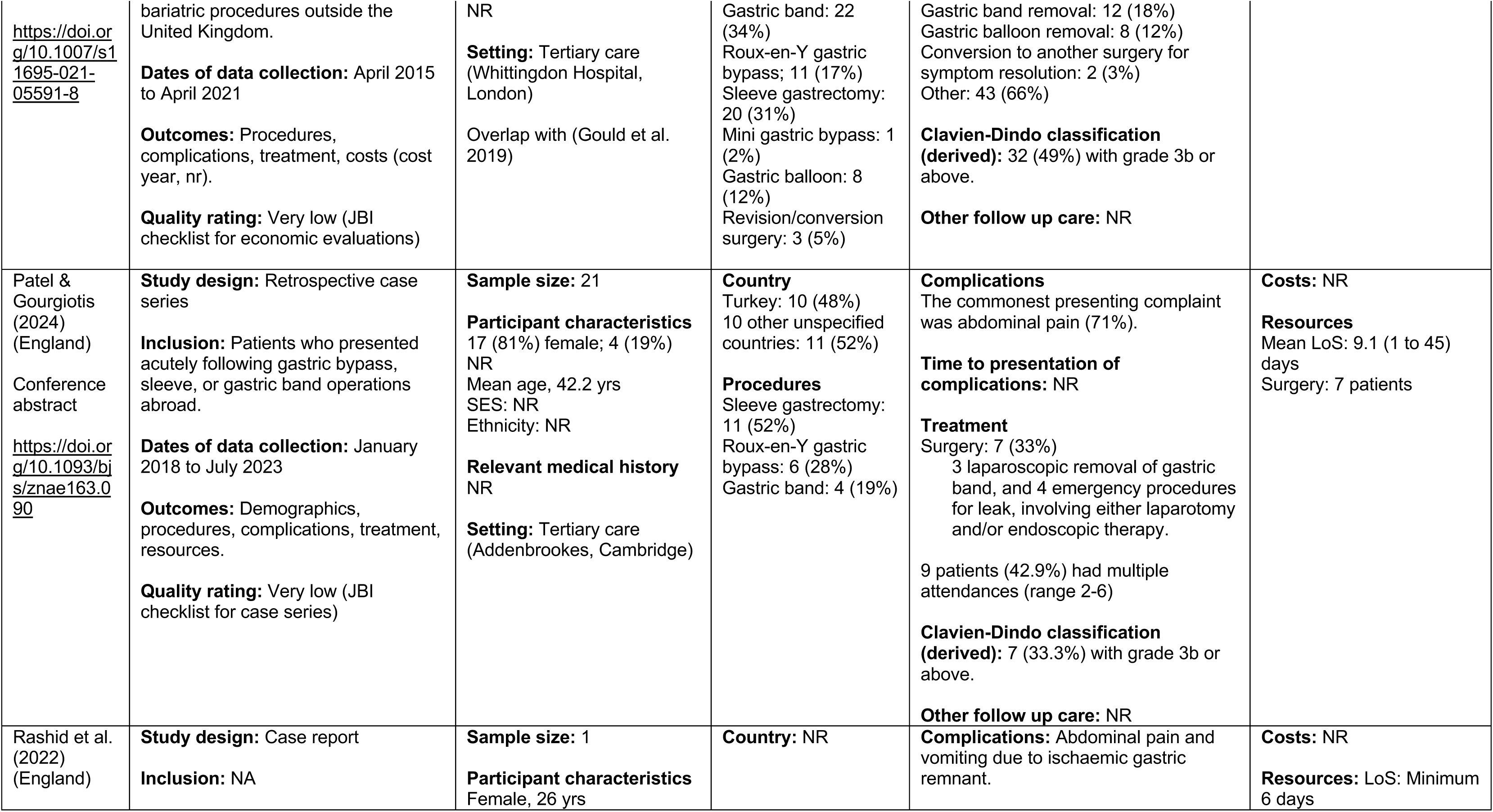
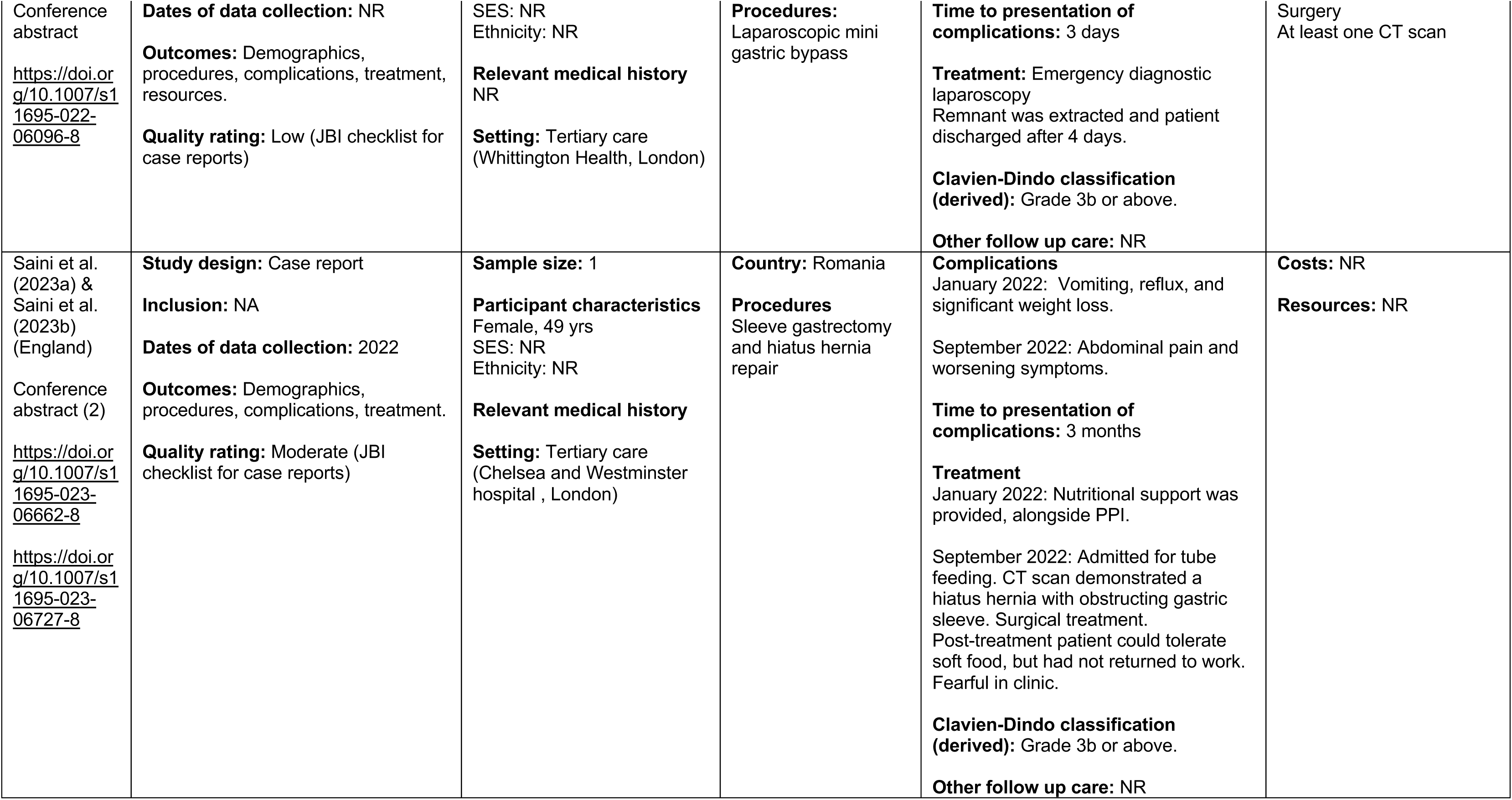
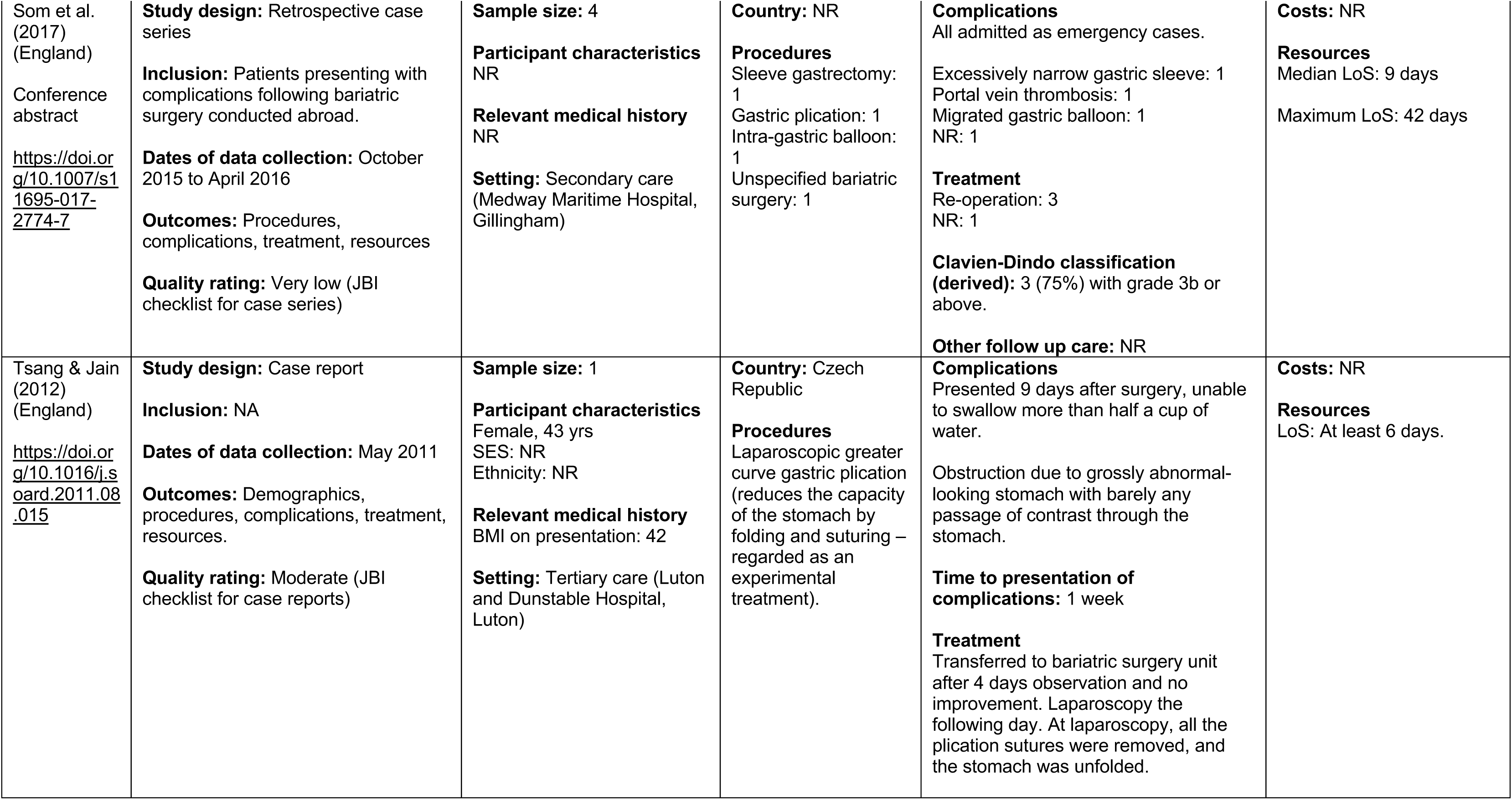
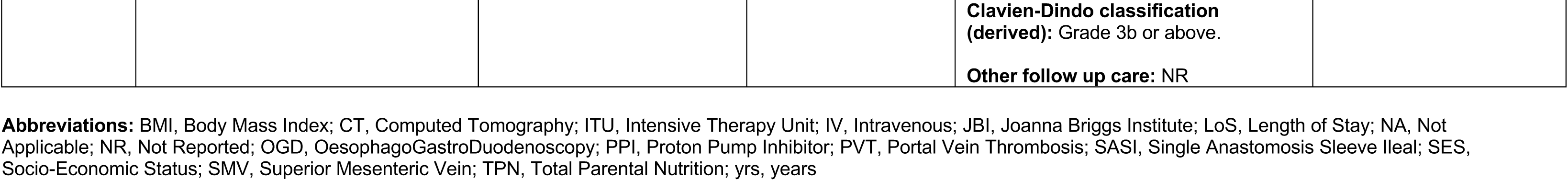
Summary of included studies: Bariatric surgery.

**Table 11:**
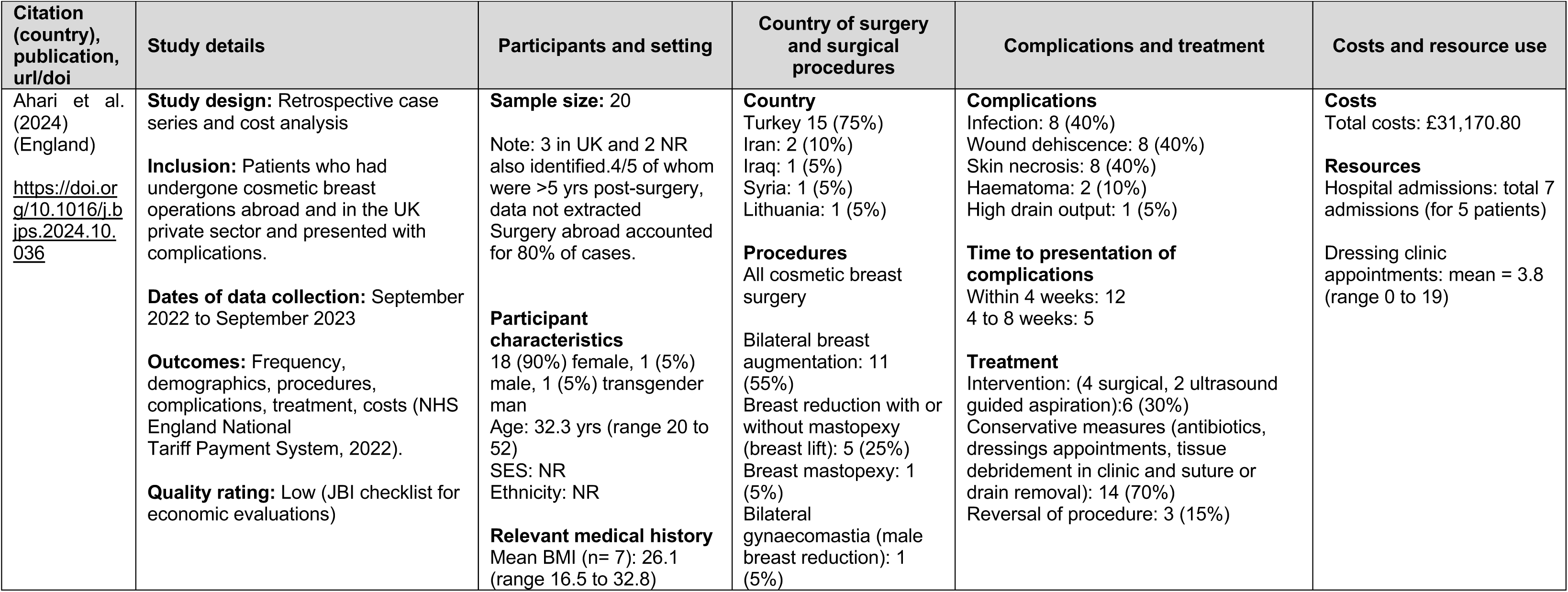
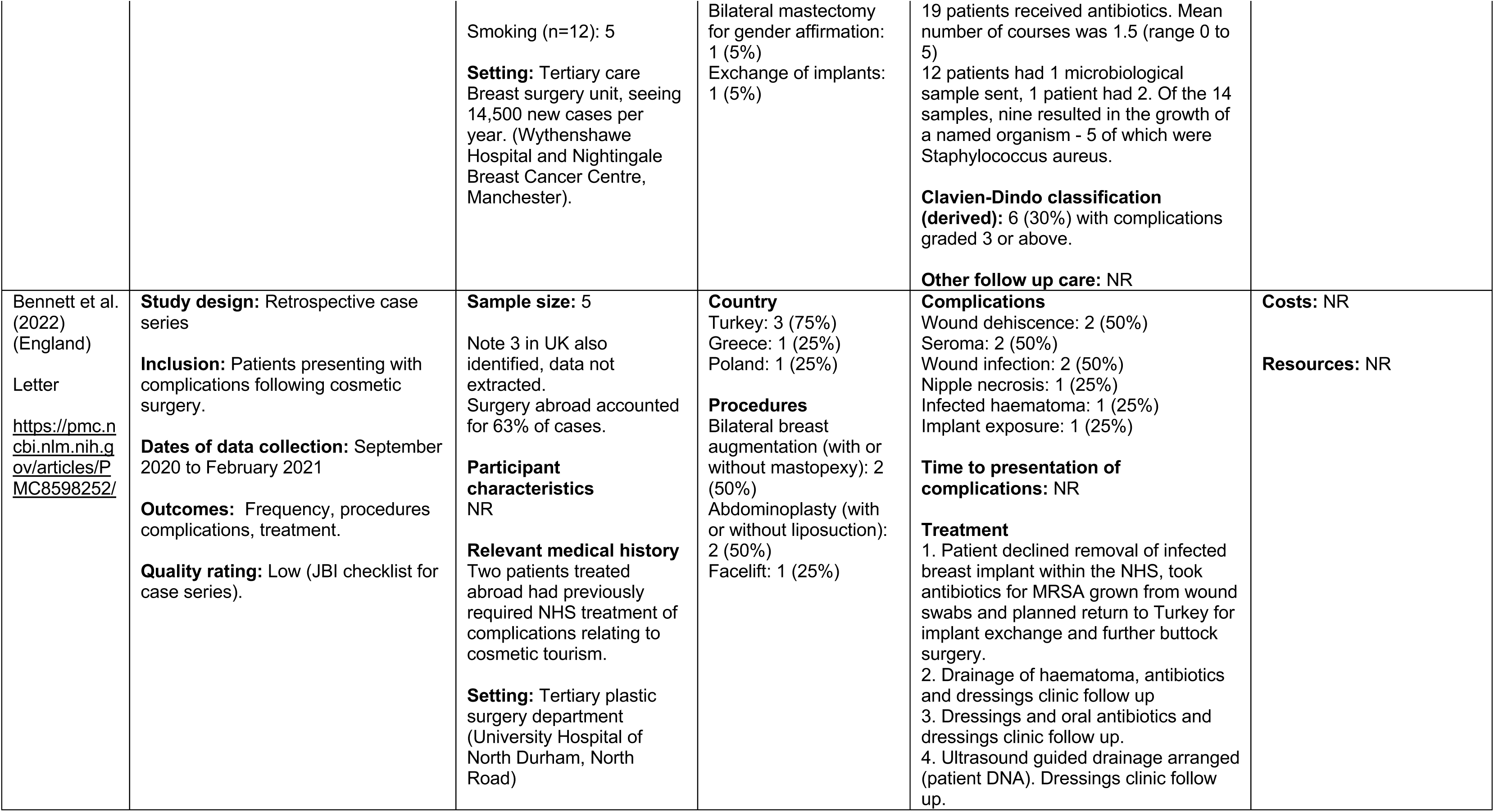
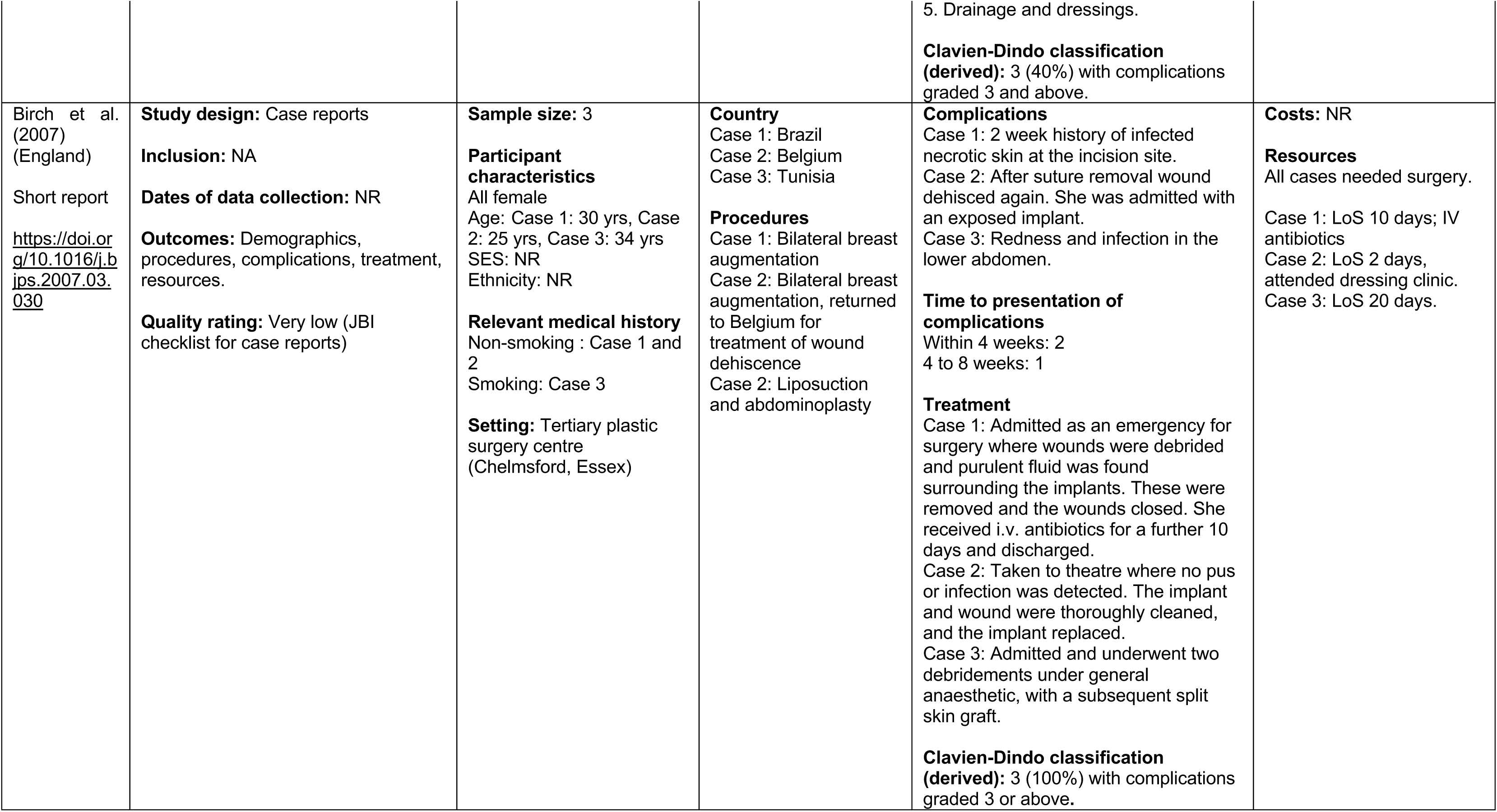
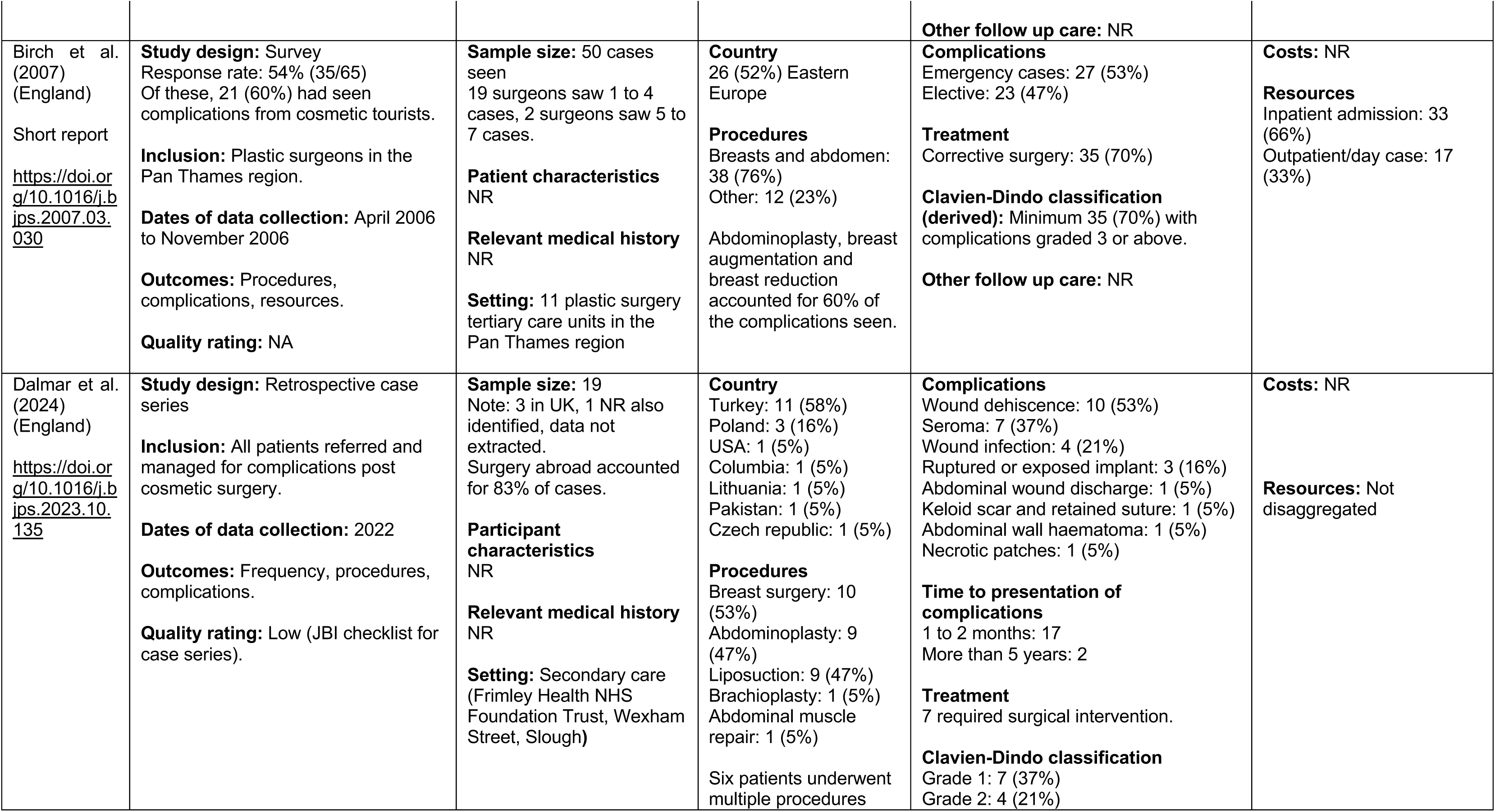
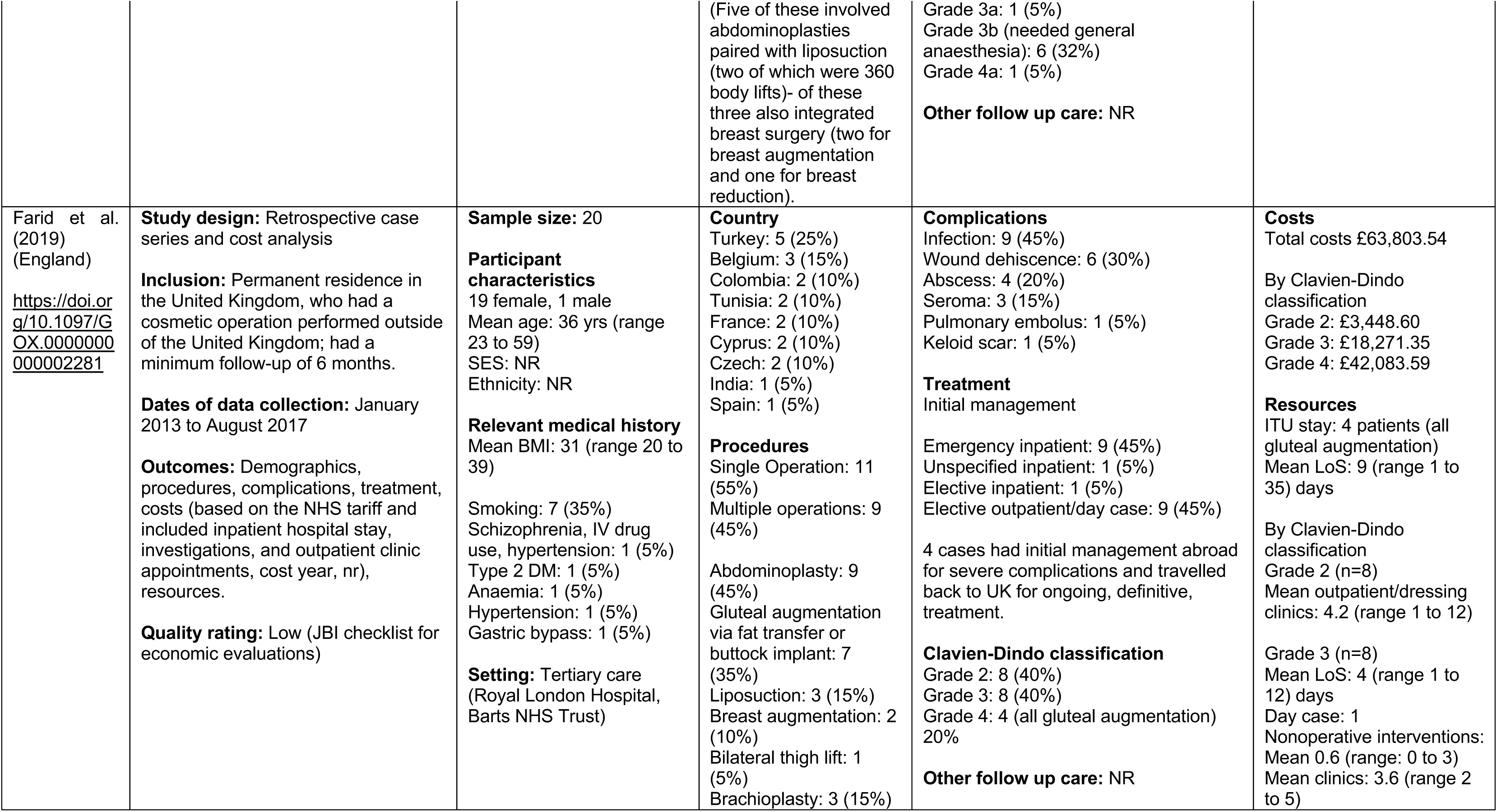
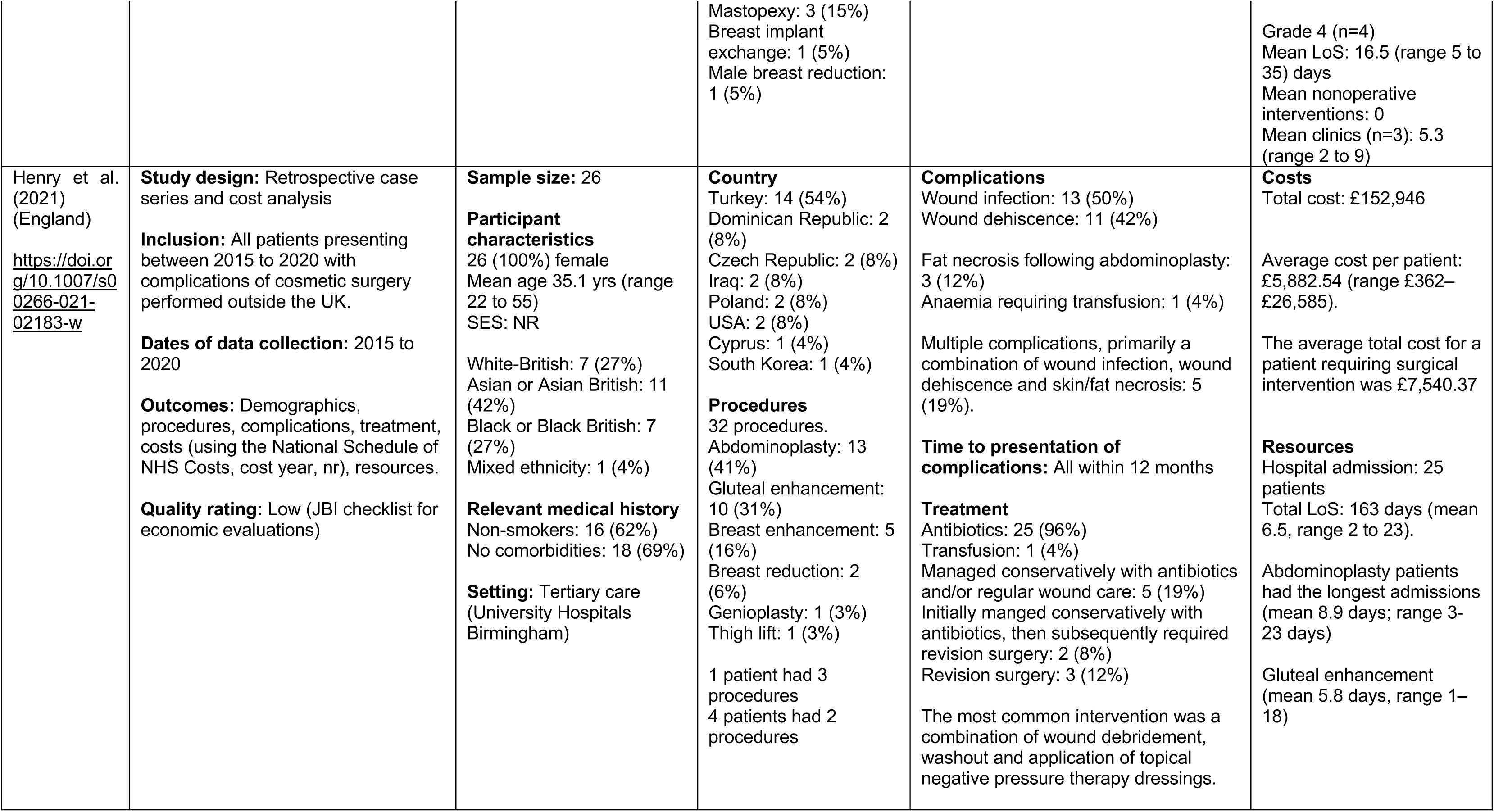
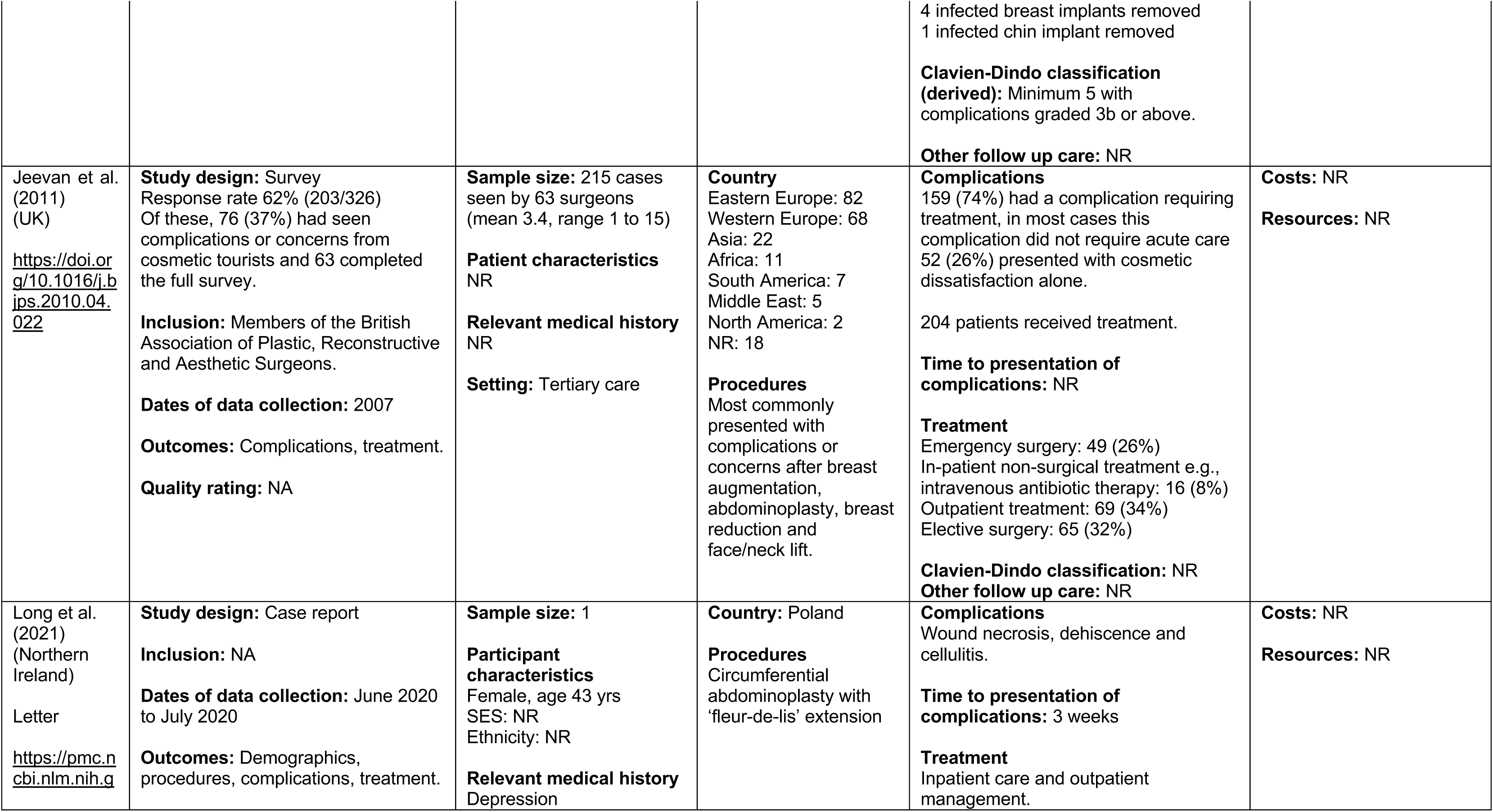
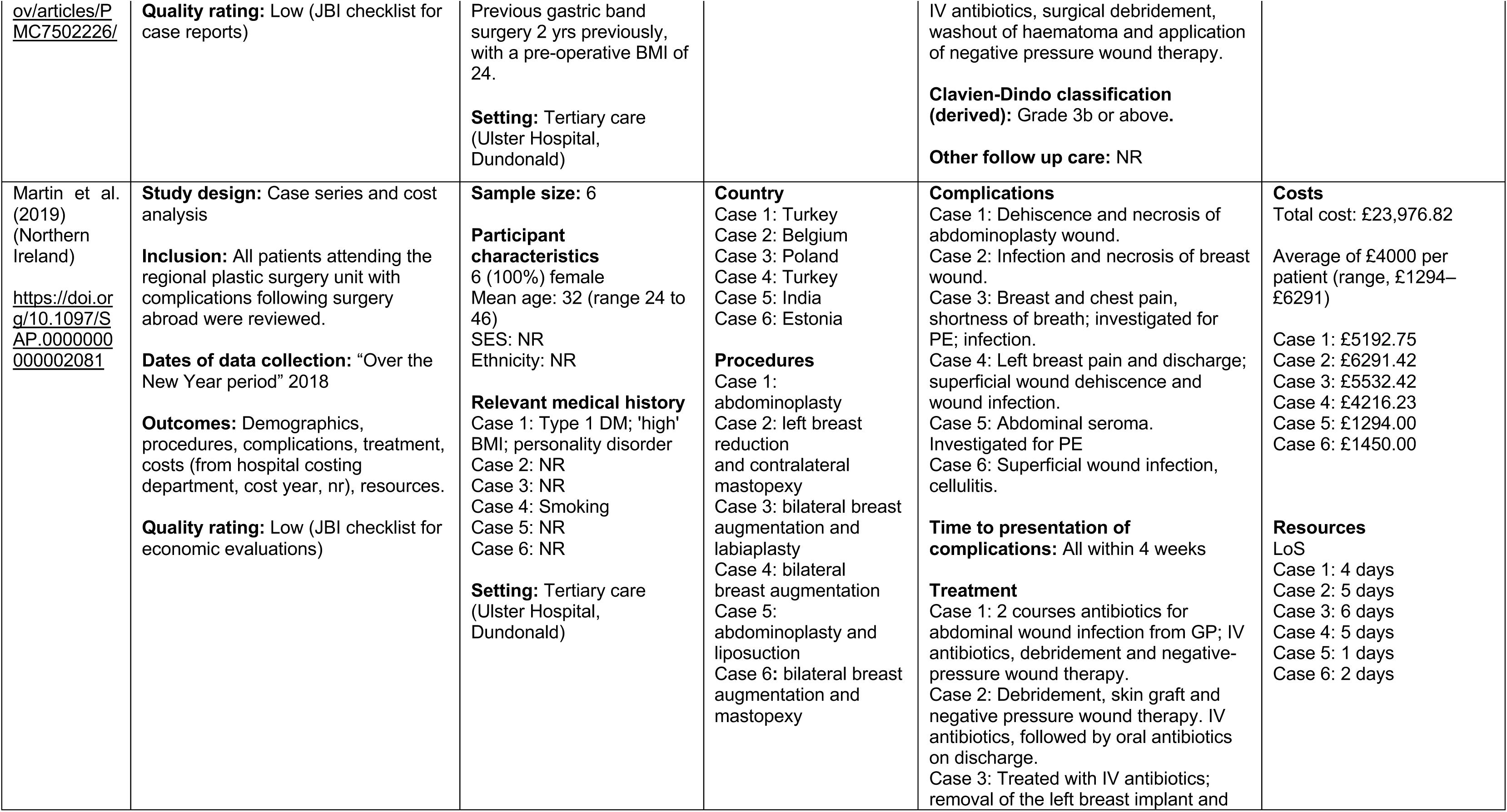
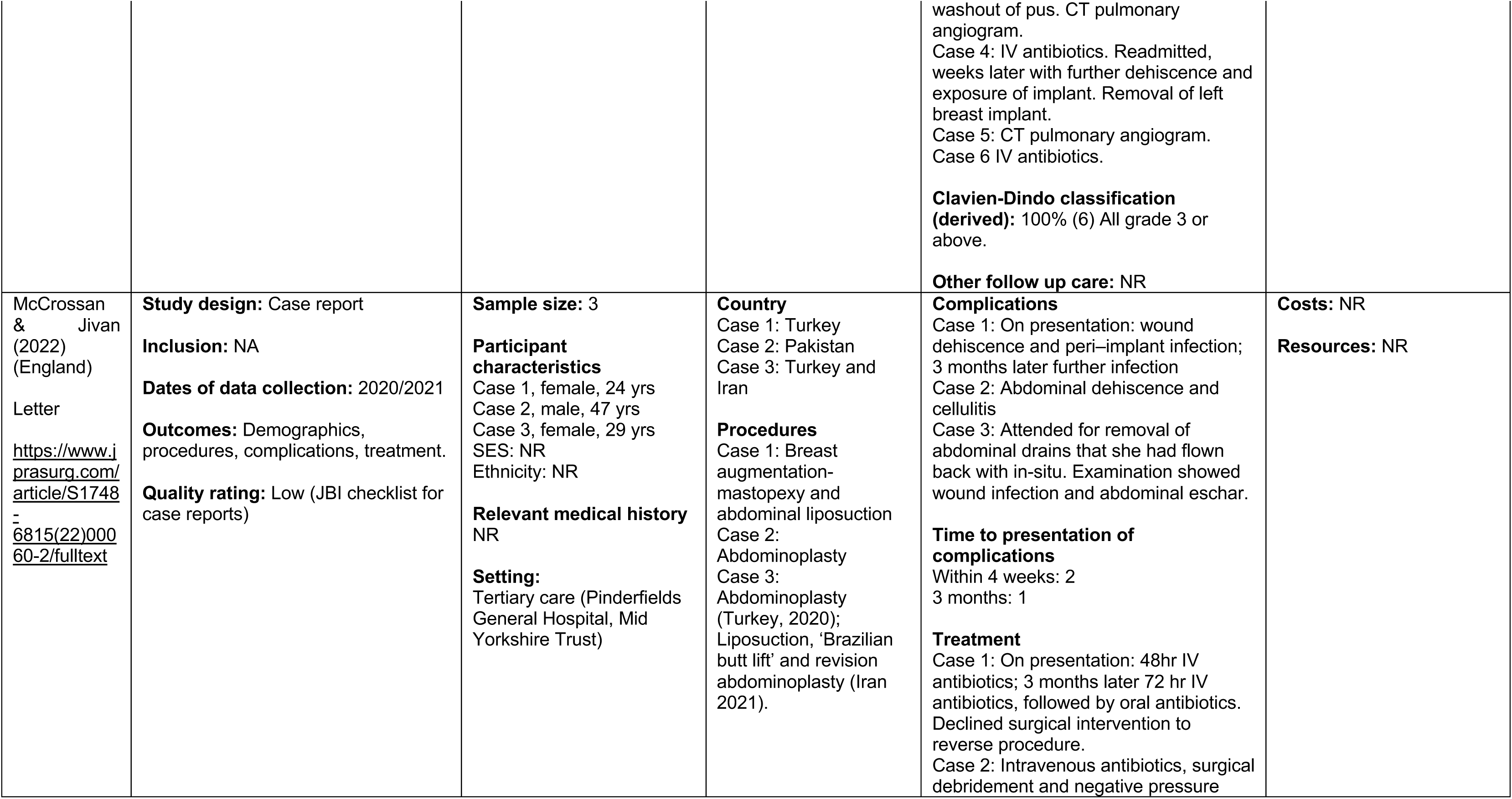
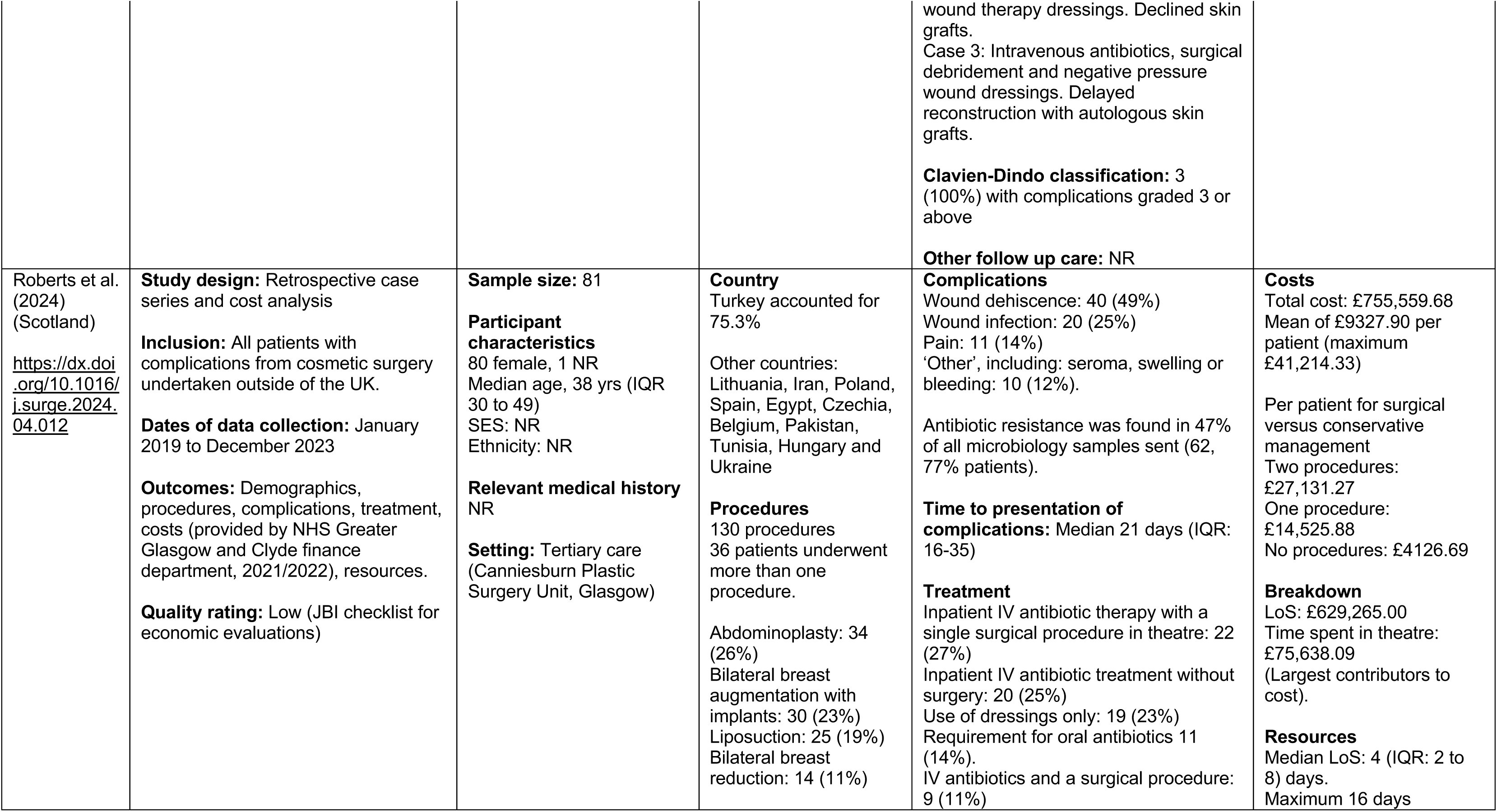
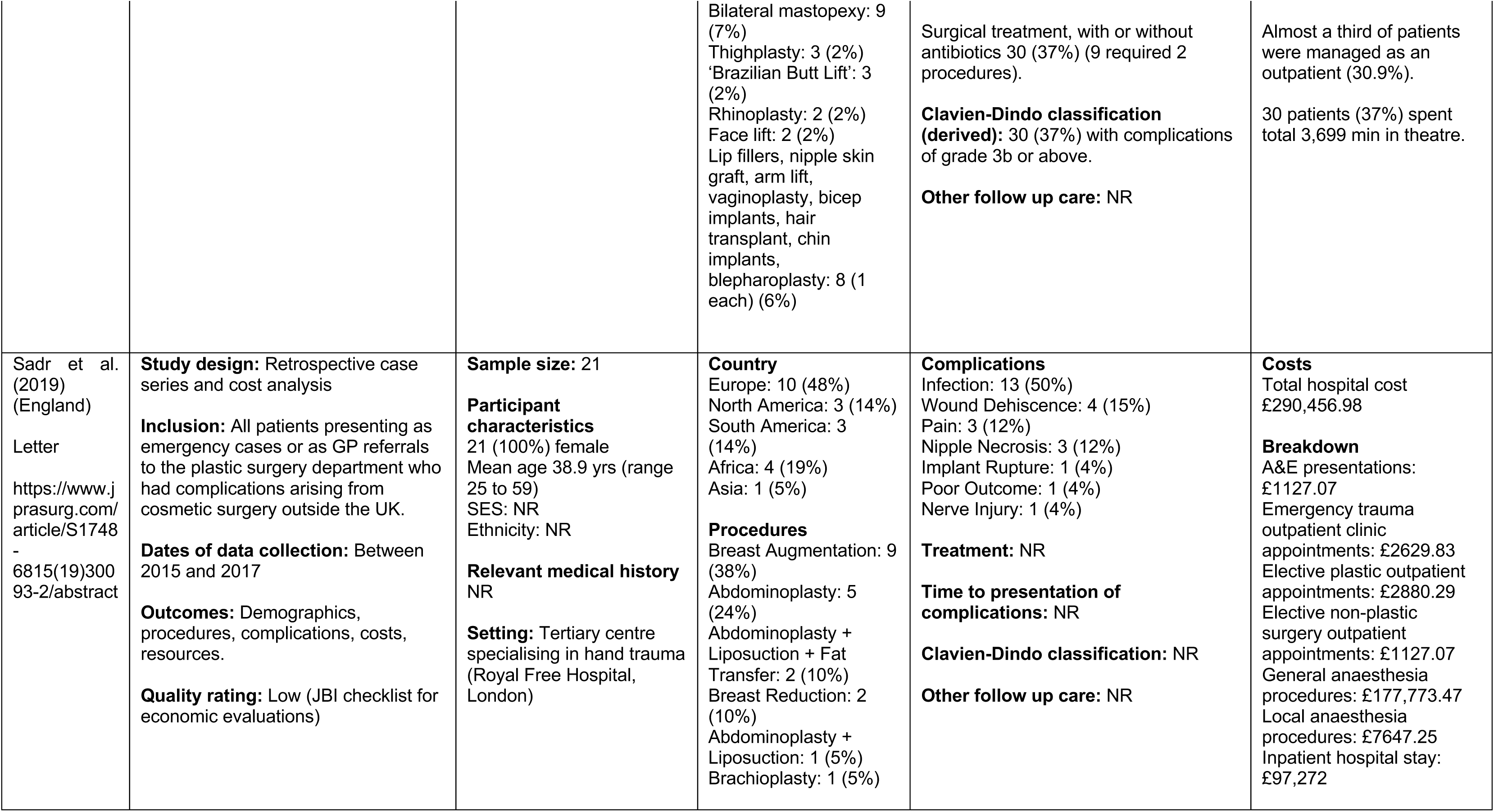
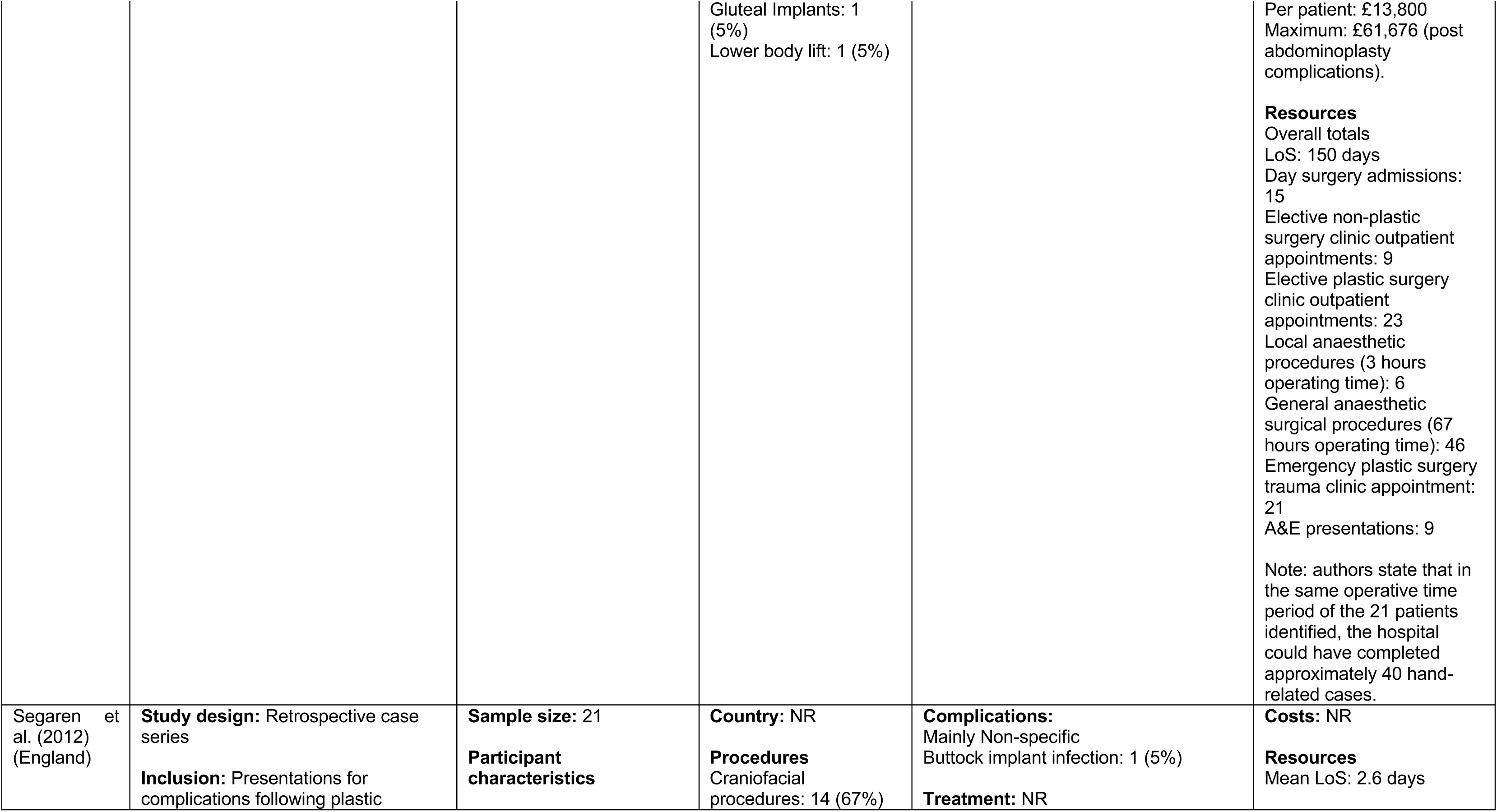
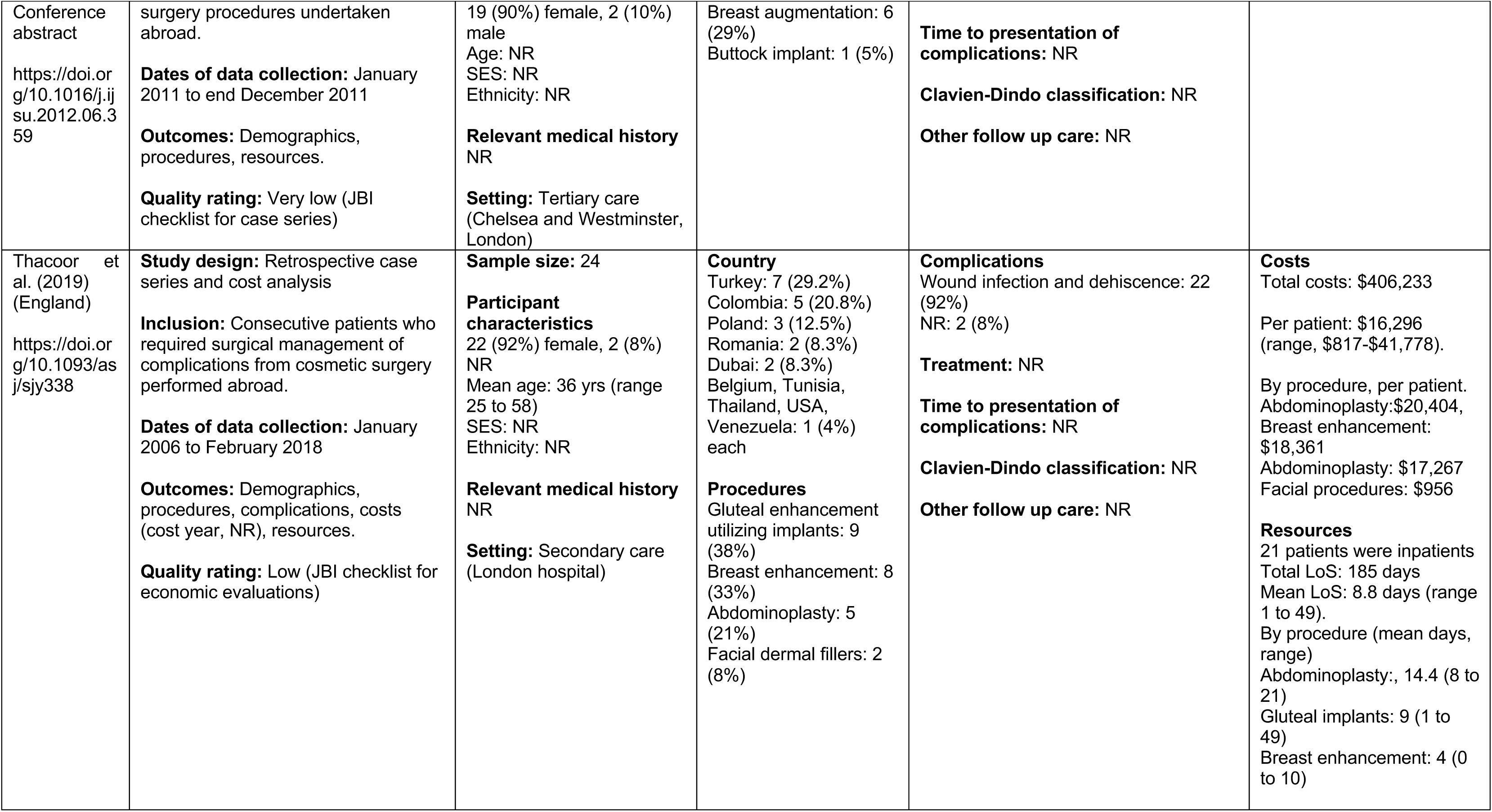
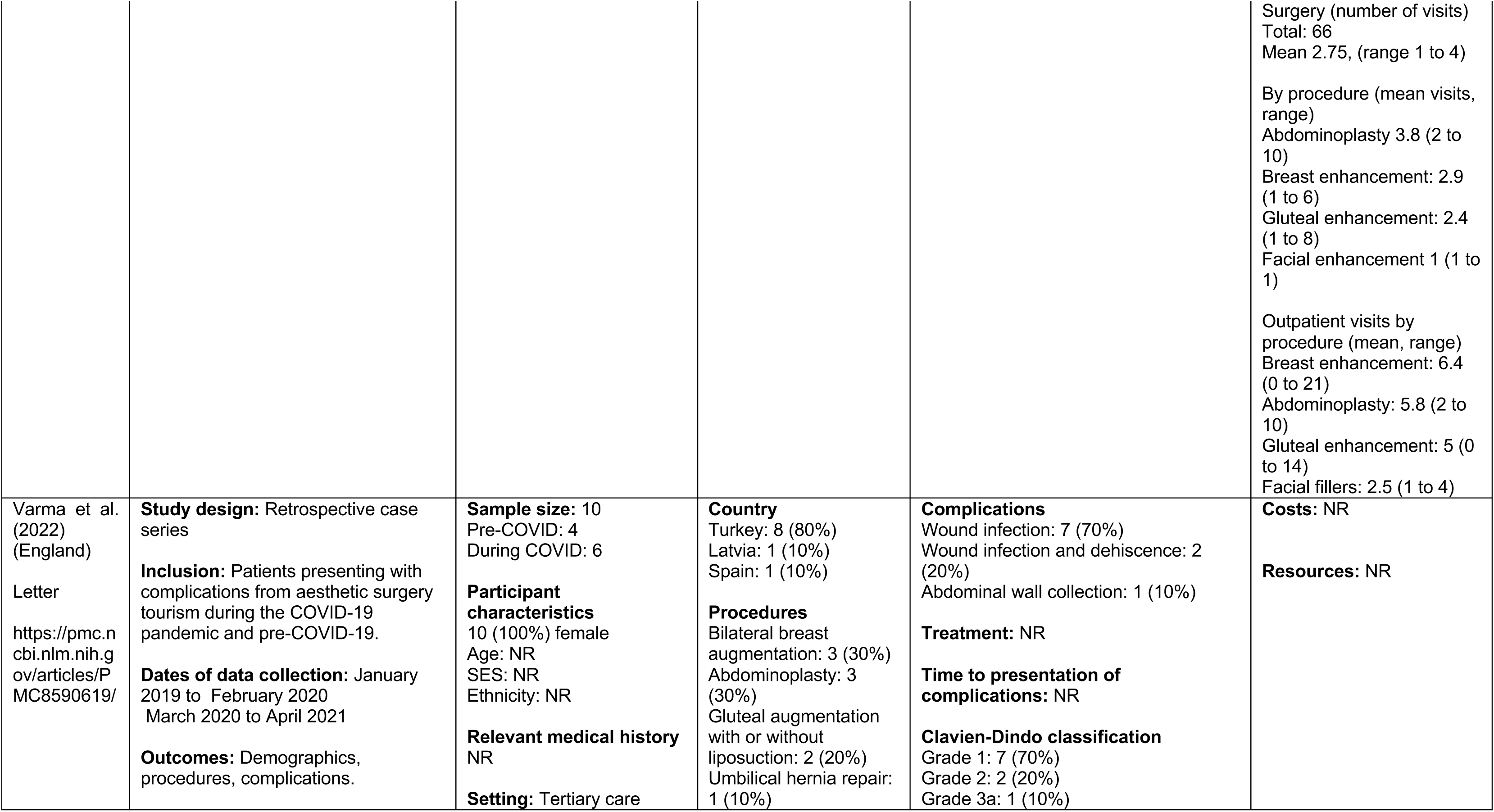
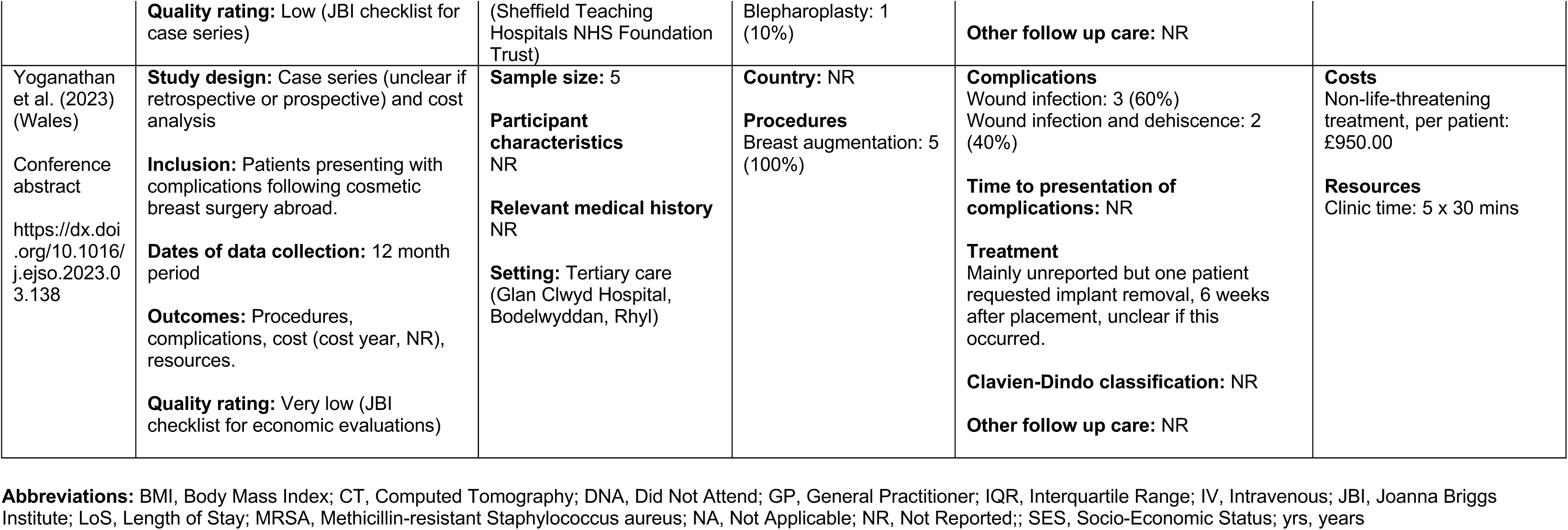
Summary of included studies: Cosmetic surgery.

**Table 12:** Summary of included studies: Ophthalmic surgery.

| Citation (country), publication, url/doi | Study details | Participants and setting | Country of surgery and surgical procedures | Complications and treatment | Costs and resource use |
| --- | --- | --- | --- | --- | --- |
| Yip et al. (2017) (England) | **Study design:** Case report<br>**Inclusion:** Cases of medical tourism seen by one surgeon over 10 years<br>**Dates of data collection:** 2006, 2011, 2015 (2) and NR | **Sample size:** 5<br>**Participant characteristics**<br>5 (100%) male<br>Case 1: 34 years<br>Case 2: 41 years | **Country**<br>Case 1, 2, 3, 4: India<br>Case 5: Panama<br>**Procedures**<br>Case 1, 2, 3: LASIK | **Complications**<br>Case 1: Reduced sight. Inflamed cornea 3 weeks post-surgery<br>Case 2: Reduced sight which deteriorated. Inflamed cornea. Developed corneal scarring and | **Costs**<br>Total: £11,583<br>Authors report this is an underestimate due to missing notes and cost of eye drops and antibiotics not included. |
| https://doi.org/10.12968/pti.2017.6.156445 | **Outcomes:** Demographics, procedures, complications, treatment, costs (NHS England 2016/17 National Prices and National Tariff Workbook), resources.<br>**Quality rating:** Very low (JBI checklist for economic evaluations) | Case 3: 51 years<br>Case 4: 35 years<br>Case 5: 29 years<br>SES: NR<br>Ethnicity: NR<br>**Relevant medical history**<br>NR<br>**Setting:** Tertiary care<br>Single surgeon | Case 4: phakic intraocular lens implant<br>Case 5: iris colour implant | superficial corneal vascularisation.<br>Case 3: Reduced sight. Fold in the flap obscuring the visual axis.<br>Case 4: Reduced sight. Staphylococcus infection 4 days post-surgery<br>Case 5: Low grade inflammation around the iris one month after surgery.<br>**Treatment**<br>Case 1: Treated with Predforte and Exocin. Post treatment visual acuity improved to 6/9 bilaterally.<br>Case 2: Initially treated with antiviral ointment, followed by topical lubricants. Then therapeutic ptosis was induced using botulinum toxin. Considered for graft but corneal scar spontaneously improved. Vision 6/9 at three years after surgery.<br>Case 3: Required two revisions and repositioning of the flap. He achieved a best corrected vision with contact lenses of 6/4 bilaterally.<br>Case 4: Intraocular lens explanation, vitrectomy and intravitreal antibiotics. His vision is now 6/60 (sight impaired) and has developed cystoid macula oedema. Right eye lens left in situ and monitored for infection. | **Resources**<br>Outpatient appointments: >50<br>Surgical procedures: 4 |
|  |  |  |  | Case 5: Topical steroid, warned of long term risks of further iritis, glaucoma and corneal decompensation. Declined removal of implants On twice-daily steroids.<br>**Clavien-Dindo classification (derived)**<br>Case 1: Grade 2<br>Case 2: Grade 2<br>Case 3: Grade 3a<br>Case 4: Grade 3a<br>Case 5: Grade 2<br>**Other follow up care:** NR |  |

**Abbreviations:** JBI, Joanna Briggs Institute; LASIK, laser-assisted in-situ-keratomileusis; NR, not reported

### 6.3 Assessment of the methodological quality of included studies

Table 13 shows the quality appraisal for cost analyses, table 14 show quality appraisal for case series and table 15 shows quality appraisal for case reports.

**Table 13.**
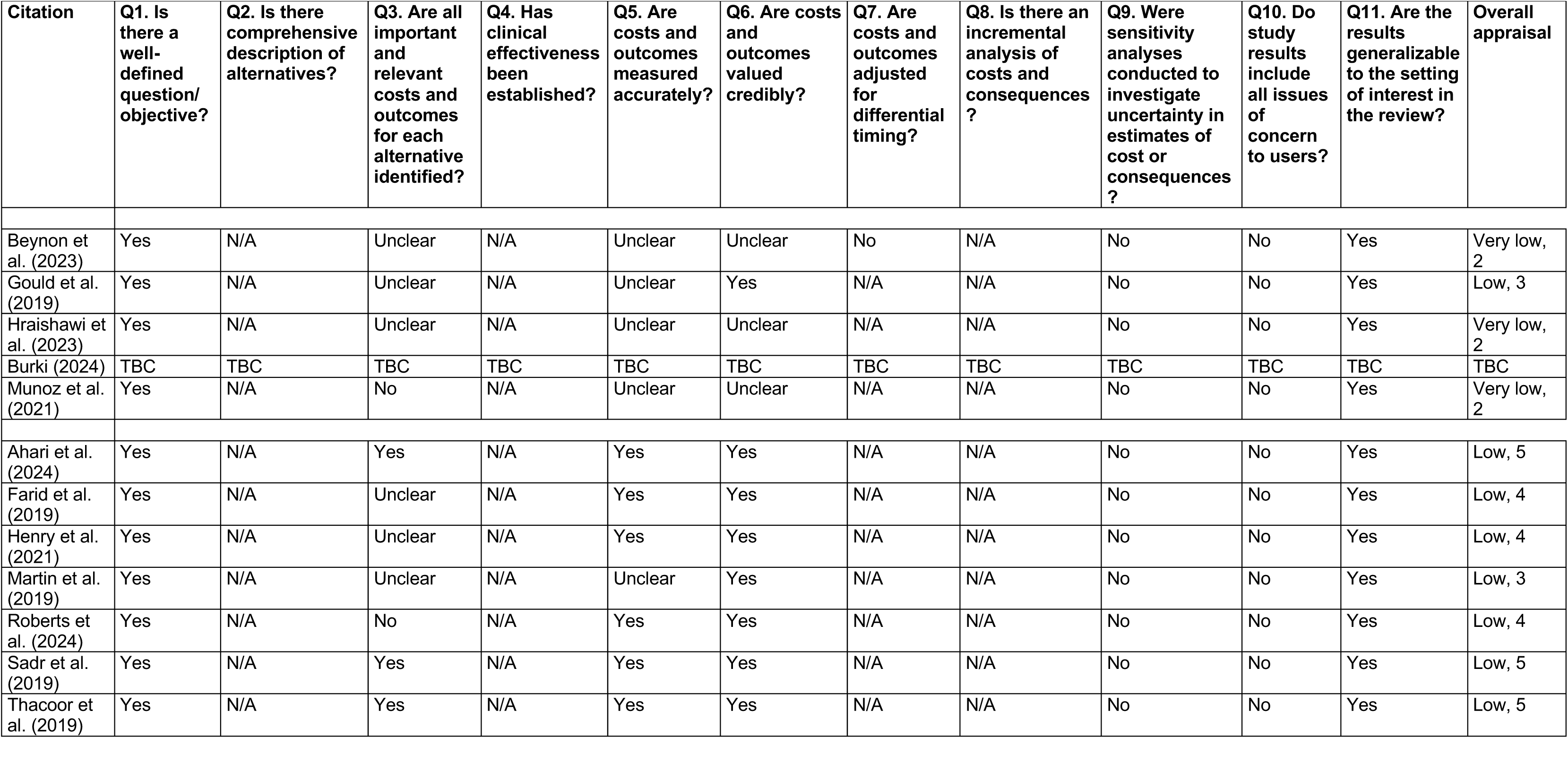
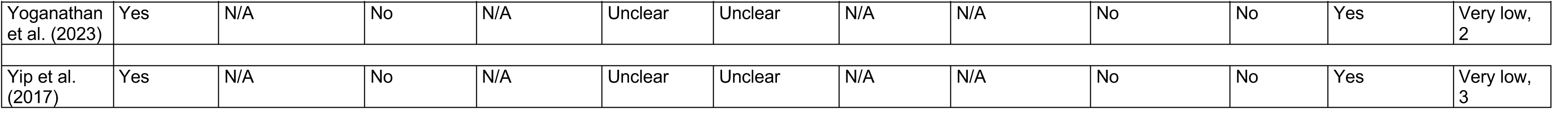
Quality appraisal for cost analyses.

| Citation | Q1. Is there a well-defined question/objective? | Q2. Is there comprehensive description of alternatives? | Q3. Are all important and relevant costs and outcomes for each alternative identified? | Q4. Has clinical effectiveness been established? | Q5. Are costs and outcomes measured accurately? | Q6. Are costs and outcomes valued credibly? | Q7. Are costs and outcomes adjusted for differential timing? | Q8. Is there an incremental analysis of costs and consequences ? | Q9. Were sensitivity analyses conducted to investigate uncertainty in estimates of cost or consequences ? | Q10. Do study results include all issues of concern to users? | Q11. Are the results generalizable to the setting of interest in the review? | Overall appraisal |
| --- | --- | --- | --- | --- | --- | --- | --- | --- | --- | --- | --- | --- |
| Beynon et al. (2023) | Yes | N/A | Unclear | N/A | Unclear | Unclear | No | N/A | No | No | Yes | Very low, 2 |
| Gould et al. (2019) | Yes | N/A | Unclear | N/A | Unclear | Yes | N/A | N/A | No | No | Yes | Low, 3 |
| Hraishawi et al. (2023) | Yes | N/A | Unclear | N/A | Unclear | Unclear | N/A | N/A | No | No | Yes | Very low, 2 |
| Burki (2024) | TBC | TBC | TBC | TBC | TBC | TBC | TBC | TBC | TBC | TBC | TBC | TBC |
| Munoz et al. (2021) | Yes | N/A | No | N/A | Unclear | Unclear | N/A | N/A | No | No | Yes | Very low, 2 |
| Ahari et al. (2024) | Yes | N/A | Yes | N/A | Yes | Yes | N/A | N/A | No | No | Yes | Low, 5 |
| Farid et al. (2019) | Yes | N/A | Unclear | N/A | Yes | Yes | N/A | N/A | No | No | Yes | Low, 4 |
| Henry et al. (2021) | Yes | N/A | Unclear | N/A | Yes | Yes | N/A | N/A | No | No | Yes | Low, 4 |
| Martin et al. (2019) | Yes | N/A | Unclear | N/A | Unclear | Yes | N/A | N/A | No | No | Yes | Low, 3 |
| Roberts et al. (2024) | Yes | N/A | No | N/A | Yes | Yes | N/A | N/A | No | No | Yes | Low, 4 |
| Sadr et al. (2019) | Yes | N/A | Yes | N/A | Yes | Yes | N/A | N/A | No | No | Yes | Low, 5 |
| Thacoor et al. (2019) | Yes | N/A | Yes | N/A | Yes | Yes | N/A | N/A | No | No | Yes | Low, 5 |

| Citation | Q1. Is there a well-defined question/objective? | Q2. Is there comprehensive description of alternatives? | Q3. Are all important and relevant costs and outcomes for each alternative identified? | Q4. Has clinical effectiveness been established? | Q5. Are costs and outcomes measured accurately? | Q6. Are costs and outcomes valued credibly? | Q7. Are costs and outcomes adjusted for differential timing? | Q8. Is there an incremental analysis of costs and consequences? | Q9. Were sensitivity analyses conducted to investigate uncertainty in estimates of cost or consequences? | Q10. Do study results include all issues of concern to users? | Q11. Are the results generalizable to the setting of interest in the review? | Overall appraisal |
| --- | --- | --- | --- | --- | --- | --- | --- | --- | --- | --- | --- | --- |
| Yoganathan et al. (2023) | Yes | N/A | No | N/A | Unclear | Unclear | N/A | N/A | No | No | Yes | Very low, 2 |
| Yip et al. (2017) | Yes | N/A | No | N/A | Unclear | Unclear | N/A | N/A | No | No | Yes | Very low, 3 |

**Table 14.** Quality appraisal for case series.

| Citation | Q1. Were there clear criteria for inclusion in the case series? | Q2. Was the condition measured in a standard, reliable way for all participants included in the case series? | Q3. Were valid methods used for identification of the condition for all participants included in the case series? | Q4. Did the case series have consecutive inclusion of participants? | Q5. Did the case series have complete inclusion of participants? | Q6. Was there clear reporting of the demographics of the participants in the study? | Q7. Was there clear reporting of clinical information of the participants? | Q8. Were the outcomes or follow up results of cases clearly reported?<br>*Note: Clarity of reporting of outcomes* | Q9. Was there clear reporting of the presenting site(s)/clinic(s) demographic information? | Q10. Was statistical analysis appropriate? | Overall appraisal |
| --- | --- | --- | --- | --- | --- | --- | --- | --- | --- | --- | --- |
| **Bariatric and cosmetic** |  |  |  |  |  |  |  |  |  |  |  |
| Beynon et al. (2023) | Yes | Unclear | Unclear | Yes | Unclear | No | No | No | No | N/A | Very low, 2 |
| **Bariatric** |  |  |  |  |  |  |  |  |  |  |  |
| Alabdellat et al. (2023) | Yes | Yes | Yes | Yes | No | No | No | No | Yes | N/A | Low, 5 |
| Alnagar et al. (2024) | Yes | Unclear | Unclear | Yes | Unclear | No | No | Yes | No | N/A | Low, 3 |
| Aly et al. (2022) | Yes | Unclear | Unclear | Yes | Unclear | No | No | No | No | N/A | Very low, 2 |
| Ftaieh et al. (2023) | Yes | Unclear | Unclear | Yes | Unclear | Partially | No | No | No | N/A | Very low, 2 |
| Gould et al. (2019) | Yes | Unclear | Unclear | Yes | Unclear | Partially (age, sex) | No | No | No | N/A | Very low, 2 |
| Hackney et al. (2022) | Yes | Unclear | Unclear | Yes | Unclear | No | No | No | No | N/A | Very low, 2 |
| Hraishawi et al. (2023) | Yes | Unclear | Unclear | Yes | Unclear | Partially (age, sex) | No | No | No | N/A | Very low, 2 |
| Iqbal et al. (2024) | Yes | Yes | Yes | Yes | No | No | No | No | Yes | N/A | Low, 4 |
| Kamel et al. (2017) | Yes | Unclear | Unclear | Yes | Yes | No | Yes | Yes | No | N/A | Low, 5 |
| Munoz et al. (2021) | Yes | Unclear | Unclear | Yes | Unclear | No | No | No | No | N/A | Very low, 2 |
| Patel & Gourgiotis (2024) | Yes | Unclear | Unclear | Yes | Unclear | Partially (age, sex) | No | No | No | N/A | Very low, 2 |
| Som et al. (2017) | Yes | Unclear | Unclear | Yes | Unclear | No | No | No | No | N/A | Very low, 2 |
| **Cosmetic** |  |  |  |  |  |  |  |  |  |  |  |
| Ahari et al. (2024) | Yes | Unclear | Unclear | Yes | Unclear | Partially (age, gender) | Yes | Yes | Yes | N/A | Low, 5 |
| Bennett et al. (2022) | Yes | Unclear | Unclear | Yes | Unclear | No | Yes | Yes | Partially | N/A | Low, 4 |
| Dalmar et al. (2024) | Yes | Unclear | Unclear | Yes | Unclear | No | Yes | No | Partially | N/A | Low, 3 |
| Farid et al. (2019) | Yes | Unclear | Unclear | Yes | No | Partially (age, gender) | Yes | Yes | No | N/A | Low, 4 |
| Henry et al. (2021) | Yes | Unclear | Unclear | Yes | Unclear | Partially (age, gender, ethnicity) | No | Yes | No | N/A | Low, 3 |
| Martin et al. (2019) | Yes | Yes | Unclear | Unclear | Unclear | Partially (age, gender) | Yes | Yes | No | N/A | Low, 4 |
| Roberts et al. (2024) | Yes | Unclear | Unclear | Yes | Unclear | Partially (age, gender) | Yes | Yes | No | N/A | Low, 4 |
| Sadr et al. (2019) | Yes | Unclear | Unclear | Yes | Unclear | Partially (age, gender) | Yes | Yes | No | N/A | Low, 4 |
| Segaren et al. (2012) | Yes | Unclear | Unclear | Yes | Unclear | No | No | No | No | N/A | Very low, 2 |
| Thacoor et al. (2019) | Yes | Unclear | Unclear | Yes | Unclear | Partially (age, gender) | No | Yes | No | N/A | Low, 3 |
| Varma et al. (2022) | Yes | Unclear | Unclear | Yes | Unclear | Partially (age) | Yes | Yes | No | N/A | Low, 4 |
| Yoganathan et al. (2023) | Yes | Unclear | Unclear | Unclear | Unclear | No | Yes | No | No | N/A | Very low, 2 |

**Table 15.** Quality appraisal for case reports.

| Citation | Q1. Were patient's demographic characteristics clearly described? | Q2. Was the patient's history clearly described and presented as a timeline? | Q3. Was the current clinical condition of the patient on presentation clearly described? | Q4. Were diagnostic tests or assessment methods and the results clearly described? | Q5. Was the intervention(s) or treatment procedure(s) clearly described? | Q6. Was the post-intervention clinical condition clearly described? | Q7. Were adverse events (harms) or unanticipated events identified and described? | Q8 Does the case report provide takeaway lessons? | Overall appraisal |
| --- | --- | --- | --- | --- | --- | --- | --- | --- | --- |
| **Bariatric** |  |  |  |  |  |  |  |  |  |
| Fakih Gomez et al. (2019) | No | No | No | No | Yes | No | Yes | Yes | Low, 3 |
| Gan et al. (2017) | No | No | Yes | Yes | Yes | No | Yes | Yes | Moderate, 5 |
| Mandour et al. (2024) | No | Yes | Yes | Yes | Yes | Yes | Yes | Yes | High, 7 |
| Rashid et al. (2022) | No | No | Yes | Yes | No | No | Yes | Yes | Low, 4 |
| Saini et al. (2023a)<br>Saini et al. (2023b) | No | No | Yes | No | Yes | Yes | Yes | Yes | Moderate, 5 |
| Tsang & Jain (2012) | No | Yes | Yes | Yes | Yes | No | Yes | Yes | Moderate, 6 |
| **Cosmetic** |  |  |  |  |  |  |  |  |  |
| Birch et al. (2007) | No | No | No | No | Yes | No | No | Yes | Very low, 2 |
| Long et al. (2021) | No | No | No | No | Yes | No | No | Yes | Low, 3 |
| McCrossan & Jivan (2022) | No | No | Yes | No | Yes | Yes | No | Yes | Low, 4 |
| **Ophthalmic** |  |  |  |  |  |  |  |  |  |
| Yip et al. (2017) | No | No | Yes | No | Yes | Yes | Yes | Yes | Moderate, 5 |

### 6.4 Assessment of the certainty of the overall body of evidence for costs

**Table 16.** Certainty of evidence for costs.

| Certainty assessment |  |  |  |  |  |  | Summary of findings |
| --- | --- | --- | --- | --- | --- | --- | --- |
| Participants (studies)<br>Follow-up | Risk of bias | Inconsistency | Indirectness | Imprecision | Publication bias | Overall certainty of evidence |  |
| **Mean cost per patient of treating complications of bariatric surgery** |  |  |  |  |  |  |  |
| 165<br>(5 non-randomised studies) | serious<sup>a</sup> | not serious | serious<sup>b</sup> | not serious | none | ⊕○○○<br>Very low<sup>a,b</sup> | Mean cost per patient ranged from £2,925 to £17,550 |
| **Mean cost per patient of treating complications of cosmetic surgery** |  |  |  |  |  |  |  |
| 203<br>(8 non-randomised studies) | serious<sup>a</sup> | not serious | serious<sup>b</sup> | not serious | none | ⊕○○○<br>Very low<sup>a,b</sup> | Mean cost per patient ranged from £950 to £13,831 |
| **Mean cost per patient of treating complications of ophthalmic surgery** |  |  |  |  |  |  |  |
| 5<br>(1 non-randomised study) | serious<sup>c</sup> | not serious | serious<sup>b</sup> | not serious | none | ⊕○○○<br>Very low<sup>b,c</sup> | Mean cost per patient was £2,317 |

CI: confidence interval

### 6.5 Information available on request

Protocol and excluded studies.

## 7. ADDITIONAL INFORMATION

### 7.2 Acknowledgements

The authors would like to thank Anna Kuczynska, Ed Wilson, Rebekah Knight and Beti-jane Ingram for their time, expertise, and contributions during stakeholder meetings in guiding the focus of the review and interpretation of findings.

## 8. APPENDIX

## APPENDIX 1: Search strategy

9. Database results (all run on the 19/11/2024)

10. Ovid MEDLINE(R) ALL

| # | Query | Results |
| --- | --- | --- |
| 1 | exp Medical Tourism/ | 1277 |
| 2 | ((health or healthcare or medical or medicine or surgical or diasporic or diaspora or cosmetic) adj3 (touris* or travel)).mp. | 5675 |
| 3 | (cross-border care or cross border care or patient care without borders or treatment* overseas or treatment* abroad or surgery abroad or surgery overseas or medical travel or travel for treatment or health treatment* abroad or medical care abroad or medical treatment* abroad or medical service trip or overseas treatment* or overseas patient*).mp. | 583 |
| 4 | ((treatment or surg* or surgical procedure* or operat*) and ("in another country" or "outside the country" or "out of the country" or "outside the UK" or "out of the UK" or abroad)).mp. | 4404 |
| 5 | international medical travel*.mp. | 16 |
| 7 | 1 or 2 or 3 or 4 or 5 | 9979 |
| 8 | limit 7 to yr="2012 -Current" | 6679 |
| 9 | exp United Kingdom/ | 400289 |
| 10 | (national health service* or nhs*).ti,ab,in. | 302854 |
| 11 | (english not ((published or publication* or translat* or written or language* or speak* or literature or citation*) adj5 english)).ti,ab. | 136157 |
| 12 | (gb or "g.b." or britain* or (british* not "british columbia") or uk or "u.k." or united kingdom* or (england* not "new england") or northern ireland* or northern irish* or scotland* or scottish* or ((wales or "south wales") not "new south wales") or welsh*).ti,ab,jw,in. | 2612125 |
| 13 | (bath or "bath's" or ((birmingham not alabama*) or ("birmingham's" not alabama*) or bradford or "bradford's" or brighton or "brighton's" or bristol or "bristol's" or carlisle* or "carlisle's" or (cambridge not (massachusetts* or boston* or harvard*)) or ("cambridge's" not (massachusetts* or boston* or harvard*)) or (canterbury not zealand*) or ("canterbury's" not zealand*) or chelmsford or "chelmsford's" or chester or "chester's" or chichester or "chichester's" or coventry or "coventry's" or derby or "derby's" or (durham not (carolina* or nc)) or ("durham's" not (carolina* or nc)) or ely or "ely's" or exeter or "exeter's" or gloucester or "gloucester's" or hereford or "hereford's" or hull or "hull's" or lancaster or "lancaster's" or leeds* or leicester or "leicester's" or (lincoln not nebraska*) or ("lincoln's" not nebraska*) or (liverpool not (new south wales* or nsw)) or ("liverpool's" not (new south wales* or nsw)) or ((london not (ontario* or ont or toronto*)) or ("london's" not (ontario* or ont or toronto*)) or manchester or "manchester's" or (newcastle not (new south wales* or nsw)) or ("newcastle's" not (new south wales* or nsw)) or norwich or "norwich's" or nottingham or "nottingham's" or oxford or "oxford's" or peterborough or "peterborough's" or plymouth or "plymouth's" or portsmouth or "portsmouth's" or preston or "preston's" or ripon or "ripon's" or salford or "salford's" or salisbury or "salisbury's" or sheffield or "sheffield's" or southampton or "southampton's" or st albans or | 1891756 |
|  | stoke or "stoke's" or sunderland or "sunderland's" or truro or "truro's" or wakefield or "wakefield's" or wells or westminster or "westminster's" or winchester or "winchester's" or wolverhampton or "wolverhampton's" or (worchester not (massachusetts* or boston* or harvard*)) or ("worchester's" not (massachusetts* or boston* or harvard*)) or (york not ("new york*" or ny or ontario* or ont or toronto*)) or ("york's" not ("new york*" or ny or ontario* or ont or toronto*))))).ti,ab,in. |  |
| 14 | (bangor or "bangor's" or cardiff or "cardiff's" or newport or "newport's" or st asaph or "st asaph's" or st davids or swansea or "swansea's").ti,ab,in. | 76735 |
| 15 | (aberdeen or "aberdeen's" or dundee or "dundee's" or edinburgh or "edinburgh's" or glasgow or "glasgow's" or inverness or (perth not australia*) or ("perth's" not australia*)) or stirling or "stirling's").ti,ab,in. | 278244 |
| 16 | (armagh or "armagh's" or belfast or "belfast's" or lisburn or "lisburn's" or londonderry or "londonderry's" or derry or "derry's" or newry or "newry's").ti,ab,in. | 37165 |
| 17 | or/9-16 | 3350888 |
| 18 | (exp africa/ or exp americas/ or exp antarctic regions/ or exp arctic regions/ or exp asia/ or exp australia/ or exp oceania/) not (exp United Kingdom/ or europe/) | 3494692 |
| 19 | 17 not 18 | 3140499 |
| 20 | 8 and 19 | 893 |

11. Embase

| # | Query | Results |
| --- | --- | --- |
| 1 | exp Medical Tourism/ | 1488 |
| 2 | ((health or healthcare or medical or medicine or surgical or diasporic or diaspora or cosmetic) adj3 (touris* or travel)).mp. | 7166 |
| 3 | (cross-border care or cross border care or patient care without borders or treatment* overseas or treatment* abroad or surgery abroad or surgery overseas or medical travel or travel for treatment or health treatment* abroad or medical care abroad or medical treatment* abroad or medical service trip or overseas treatment* or overseas patient*).mp. | 819 |
| 4 | ((treatment or surg* or surgical procedure* or operat*) and ("in another country" or "outside the country" or "out of the country" or "outside the UK" or "out of the UK" or abroad)).mp. | 8113 |
| 5 | international medical travel*.mp. | 19 |
| 6 | 1 or 2 or 3 or 4 or 5 | 15254 |
| 8 | limit 7 to yr="2012 -Current" | 10369 |
| 9 | exp United Kingdom/ | 484036 |
| 10 | (national health service* or nhs*).ti,ab,in. | 447157 |
| 11 | (english not ((published or publication* or translat* or written or language* or speak* or literature or citation*) adj5 english)).ti,ab. | 67506 |
| 12 | (gb or "g.b." or britain* or (british* not "british columbia") or uk or "u.k." or united kingdom* or (england* not "new england") or northern ireland* or northern irish* or scotland* or scottish* or ((wales or "south wales") not "new south wales") or welsh*).ti,ab,jw,in. | 3852272 |
| 13 | (bath or "bath's" or ((birmingham not alabama*) or ("birmingham's" not alabama*)) or bradford or "bradford's" or brighton or "brighton's" or bristol or "bristol's" or carlisle* or "carlisle's" or (cambridge not (massachusetts* or | 3064508 |
|  | boston* or harvard*) or ("cambridge's" not (massachusetts* or boston* or harvard*)) or (canterbury not zealand*) or ("canterbury's" not zealand*) or chelmsford or "chelmsford's" or chester or "chester's" or chichester or "chichester's" or coventry or "coventry's" or derby or "derby's" or (durham not (carolina* or nc)) or ("durham's" not (carolina* or nc)) or ely or "ely's" or exeter or "exeter's" or gloucester or "gloucester's" or hereford or "hereford's" or hull or "hull's" or lancaster or "lancaster's" or leeds* or leicester or "leicester's" or (lincoln not nebraska*) or ("lincoln's" not nebraska*) or (liverpool not (new south wales* or nsw)) or ("liverpool's" not (new south wales* or nsw)) or ((london not (ontario* or ont or toronto*)) or ("london's" not (ontario* or ont or toronto*)) or manchester or "manchester's" or (newcastle not (new south wales* or nsw)) or ("newcastle's" not (new south wales* or nsw)) or norwich or "norwich's" or nottingham or "nottingham's" or oxford or "oxford's" or peterborough or "peterborough's" or plymouth or "plymouth's" or portsmouth or "portsmouth's" or preston or "preston's" or ripon or "ripon's" or salford or "salford's" or salisbury or "salisbury's" or sheffield or "sheffield's" or southampton or "southampton's" or st albans or stoke or "stoke's" or sunderland or "sunderland's" or truro or "truro's" or wakefield or "wakefield's" or wells or westminster or "westminster's" or winchester or "winchester's" or wolverhampton or "wolverhampton's" or (worchester not (massachusetts* or boston* or harvard*)) or ("worchester's" not (massachusetts* or boston* or harvard*)) or (york not ("new york*" or ny or ontario* or ont or toronto*)) or ("york's" not ("new york*" or ny or ontario* or ont or toronto*))))).ti,ab,in. |  |
| 14 | (bangor or "bangor's" or cardiff or "cardiff's" or newport or "newport's" or st asaph or "st asaph's" or st davids or swansea or "swansea's").ti,ab,in. | 126567 |
| 15 | (aberdeen or "aberdeen's" or dundee or "dundee's" or edinburgh or "edinburgh's" or glasgow or "glasgow's" or inverness or (perth not australia*) or ("perth's" not australia*) or stirling or "stirling's").ti,ab,in. | 422530 |
| 16 | (armagh or "armagh's" or belfast or "belfast's" or lisburn or "lisburn's" or londonderry or "londonderry's" or derry or "derry's" or newry or "newry's").ti,ab,in. | 59334 |
| 17 | or/9-16 | 4718666 |
| 18 | (exp africa/ or exp americas/ or exp antarctic regions/ or exp arctic regions/ or exp asia/ or exp australia/ or exp oceania/) not (exp United Kingdom/ or europe/) | 3856709 |
| 19 | 17 not 18 | 4436674 |
| 20 | 8 and 19 | 1593 |

12. CINAHL Plus with Full Text Interface - EBSCOhost Research Databases:

| # | Query | Results |
| --- | --- | --- |
| S9 | S1 OR S2 OR S3 OR S4 OR S5 OR S6<br>Narrow by SubjectGeographic: - uk & ireland<br>Search modes - Proximity | 442 |
| S8 | S1 OR S2 OR S3 OR S4 OR S5 OR S6<br>Limiters - Publication Date: 20120101-20241231 | 1,648 |
| S7 | S1 OR S2 OR S3 OR S4 OR S5 | 2,232 |
| S5 | TI "international medical travel*" OR AB "international medical travel" | 8 |
| S4 | TI ( ((treatment or surg* or operat*) and ("in another country" or "outside the country" or "out of the country" or "outside the UK" or "out of the UK" or abroad)) ) OR AB ( ((treatment or surg* or surgical procedure* or operat*) and ("in another country" or "outside the country" or "out of the country" or "outside the UK" or "out of the UK" or abroad)) ) | 977 |
| S3 | TI ("cross-border care" OR "cross border care" OR "patient care without borders" OR "treatment* overseas" OR "treatment* abroad" OR "surgery abroad" OR "surgery overseas" OR "medical travel" OR "travel for treatment" OR "health treatment* abroad" OR "medical care abroad" OR "medical treatment* abroad" OR "medical service trip" OR "overseas treatment*" OR "overseas patient*")) OR AB ("cross-border care" OR "cross border care" OR "patient care without borders" OR "treatment* overseas" OR "treatment* abroad" OR "surgery abroad" OR "surgery overseas" OR "medical travel" OR "travel for treatment" OR "health treatment* abroad" OR "medical care abroad" OR "medical treatment* abroad" OR "medical service trip" OR "overseas treatment*" OR "overseas patient*")) | 253 |
| S2 | TI ( ((health or healthcare or medical or medicine or surgical or diasporic or diaspora or cosmetic) W2 (touris* or travel)) ) OR AB ( ((health or healthcare or medical or medicine or surgical or diasporic or diaspora or cosmetic) W2 (touris* or travel)) ) | 1,086 |
| S1 | (MM "Medical Tourism") | 404 |

13. Scopus

| # | Query | Results |
| --- | --- | --- |
| 7 | ( TITLE-ABS-KEY ( "international medical travel*" ) ) OR ( TITLE-ABS-KEY ( ( treatment OR surg* OR operat* ) AND ( "in another country" OR "outside the country" OR "out of the country" OR "outside the UK" OR "out of the UK" OR abroad ) ) ) OR ( TITLE-ABS-KEY ( ( "cross-border care" OR "cross border care" OR "patient care without borders" OR "treatment* overseas" OR "treatment* abroad" OR "surgery abroad" OR "surgery overseas" OR "medical travel" OR "travel for treatment" OR "health treatment* abroad" OR "medical care abroad" OR "medical treatment* abroad" OR "medical service trip" OR "overseas treatment*" OR "overseas patient*" ) ) ) OR ( TITLE-ABS-KEY ( ( health OR healthcare OR medical OR medicine OR surgical OR diasporic OR diaspora OR cosmetic ) W/2 ( touris* OR travel ) ) ) AND PUBYEAR > 2011 AND PUBYEAR < 2026 AND ( LIMIT-TO ( AFFILCOUNTRY , "United Kingdom" ) ) | 1,434 |
| 6 | ( TITLE-ABS-KEY ( "international medical travel*" ) ) OR ( TITLE-ABS-KEY ( ( treatment OR surg* OR operat* ) AND ( "in another country" OR "outside the country" OR "out of the country" OR "outside the UK" OR "out of the UK" OR abroad ) ) ) OR ( TITLE-ABS-KEY ( ( "cross-border care" OR "cross border care" OR "patient care without borders" OR "treatment* overseas" OR "treatment* abroad" OR "surgery abroad" OR "surgery overseas" OR "medical travel" OR "travel for treatment" OR "health treatment* abroad" OR "medical care abroad" OR "medical treatment* abroad" OR "medical service trip" OR "overseas treatment*" OR "overseas patient*" ) ) ) OR ( TITLE-ABS-KEY ( ( health OR healthcare OR medical OR medicine OR surgical OR diasporic OR diaspora OR cosmetic ) W/2 ( touris* OR travel ) ) ) AND PUBYEAR > 2011 AND PUBYEAR < 2026 | 19,585 |
| 5 | ( TITLE-ABS-KEY ( "international medical travel*" ) ) OR ( TITLE-ABS-KEY ( ( treatment OR surg* OR operat* ) AND ( "in another country" OR "outside the country" OR "out of the country" OR "outside the UK" OR "out of the UK" OR abroad ) ) ) OR ( TITLE-ABS-KEY ( ( "cross-border care" OR "cross border care" OR "patient care without borders" OR "treatment* overseas" OR "treatment* abroad" OR "surgery abroad" OR "surgery overseas" OR "medical travel" OR "travel for treatment" OR "health treatment* abroad" OR "medical care abroad" OR "medical treatment* abroad" OR "medical service trip" OR "overseas treatment*" OR "overseas patient*" ) ) ) OR ( TITLE-ABS-KEY ( ( health OR healthcare OR medical OR medicine OR surgical OR diasporic OR diaspora OR cosmetic ) W/2 ( touris* OR travel ) ) ) | 28,058 |
| 4 | TITLE-ABS-KEY ( "international medical travel*" ) | 51 |
| 3 | TITLE-ABS-KEY ( ( treatment OR surg* OR operat* ) AND ( "in another country" OR "outside the country" OR "out of the country" OR "outside the UK" OR "out of the UK" OR abroad ) ) | 16,406 |
| 2 | TITLE-ABS-KEY ( ( "cross-border care" OR "cross border care" OR "patient care without borders" OR "treatment* overseas" OR "treatment* abroad" OR "surgery abroad" OR "surgery overseas" OR "medical travel" OR "travel for treatment" OR "health treatment* abroad" OR "medical care abroad" OR "medical treatment* abroad" OR "medical service trip" OR "overseas treatment*" OR "overseas patient*" ) ) | 919 |
| 1 | TITLE-ABS-KEY ( ( health OR healthcare OR medical OR medicine OR surgical OR diasporic OR diaspora OR cosmetic ) W/2 ( touris* OR travel ) ) | 11,870 |

14. ProQuest Dissertations & Theses Global

| # | Query | Results |
| --- | --- | --- |
| S1 | subject(medical tourism) | 177 |
| S2 | title(((health OR healthcare OR medical OR medicine OR surgical OR diasporic OR diaspora OR cosmetic) NEAR/2 (touris* OR travel))) OR abstract(((health OR healthcare OR medical OR medicine OR surgical OR diasporic OR diaspora OR cosmetic) NEAR/2 (touris* OR travel))) | 671 |
| S3 | title(("cross-border care" OR "cross border care" OR "patient care without borders" OR "treatment* overseas" OR "treatment* abroad" OR "surgery abroad" OR "surgery overseas" OR "medical travel" OR "travel for treatment" OR "health treatment* abroad" OR "medical care abroad" OR "medical treatment* abroad" OR "medical service trip" OR "overseas treatment*" OR "overseas patient*")) OR abstract(("cross-border care" OR "cross border care" OR "patient care without borders" OR "treatment* overseas" OR "treatment* abroad" OR "surgery abroad" OR "surgery overseas" OR "medical travel" OR "travel for treatment" OR "health treatment* abroad" OR "medical care abroad" OR "medical treatment* abroad" OR "medical service trip" OR "overseas treatment*" OR "overseas patient*")) | 38 |
| S4 | title((treatment OR surg* OR operat*) AND ("in another country" OR "outside the country" OR "out of the country" OR "outside the UK" OR "out of the UK" OR abroad)) OR abstract((treatment OR surg* OR operat*) AND ("in another country" OR "outside the country" OR "out of the country" OR "outside the UK" OR "out of the UK" OR abroad)) | 9226 |
| S5 | title(international medical travel*) OR abstract(international medical travel*) | 5 |
| S6 | [S1] OR [S2] OR [S3] OR [S4] OR [S5] | 9958 |
| S7 | [S1] OR [S2] OR [S3] OR [S4] OR [S5]<br>Narrowed by:<br>Entered date: 2012-01-01 - 2024-11-19 | 1473 |
| S8 | [S1] OR [S2] OR [S3] OR [S4] OR [S5]<br>Narrowed by: University/institution location: England; Scotland; Wales; Northern Ireland;<br>Entered date: 2012-01-01 - 2024-11-19 | 145 |

15. Cochrane Library

| # | Query | Results |
| --- | --- | --- |
| #1 | MeSH descriptor: [Medical Tourism] explode all trees | 2 |
| #2 | ((health or healthcare or medical or medicine or surgical or diasporic or diaspora or cosmetic) NEAR/3 (touris* or travel)):ti,ab,kw | 237 |
| #3 | ("cross-border care" OR "cross border care" OR "patient care without borders" OR (treatment* NEXT overseas) OR (treatment* NEXT abroad) OR "surgery abroad" OR "surgery overseas" OR "medical travel" OR "travel for treatment" OR (health treatment* NEXT abroad) OR "medical care abroad" OR (medical treatment* NEXT abroad) OR "medical service trip" OR "overseas treatment*" OR "overseas patient*"):ti,ab,kw | 9 |
| #4 | ((treatment OR surg* OR operat* ) AND ( "in another country" OR "outside the country" OR "out of the country" OR "outside the UK" OR "out of the UK" OR abroad )):ti,ab,kw | 284 |
| #5 | ("international medical" NEXT travel*):ti,ab,kw | 0 |
| #6 | (OR #1 #5) | 526 |
| #7 | (OR #1 #5) with Cochrane Library publication date Between Jan 2012 and Dec 2024 | 487 |

NB. 1 review, 486 trials. Not Limited to the UK

16. Summary of results

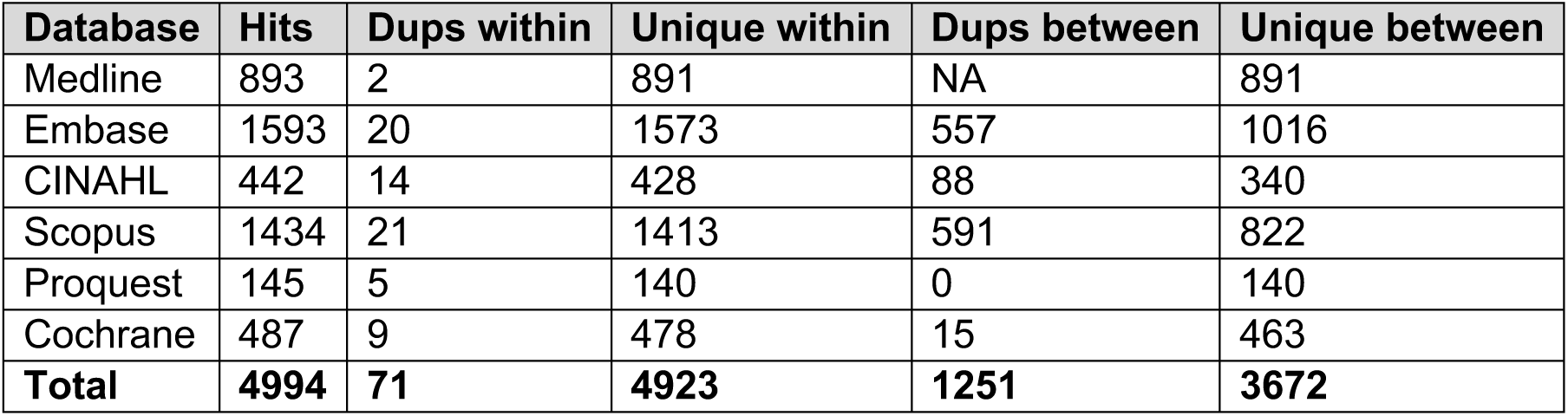

